# Predicting Phenoconversion to Clinically Manifest ALS: Results of a Large-Scale Proteomic Study

**DOI:** 10.64898/2025.12.06.25341403

**Authors:** Ximing Ran, Joanne Wuu, Zhaohui S. Qin, Johnathan Cooper-Knock, Volkan Granit, Anne-Laure Grignon, Yindi Li, Eliza Lin, Maria Catalina Fernandez, Danielle Colato, Nathan Carberry, Christina M. Lill, Paolo Piazza, Andrea Malaspina, Michael Benatar

**Affiliations:** Department of Biostatistics and Bioinformatics, Emory University, Atlanta, GA, USA; Department of Neurology and ALS Center, University of Miami Miller School of Medicine, Miami, FL, USA; Sheffield Institute for Translational Neuroscience (SITraN), University of Sheffield, Sheffield, UK; NIHR Sheffield Biomedical Research Centre, Sheffield, UK; UCL Queen Square Motor Neuron Disease Center, UCL Queen Square Institute of Neurology, University College London, Queen Square, London, UK; Institute of Epidemiology and Social Medicine, University of Münster, Münster, Germany; Ageing and Epidemiology Unit (AGE), School of Public Health, Imperial College London, London, UK; Nuffield Department of Medicine, Centre for Human Genetics, University of Oxford, Oxford, UK

## Abstract

The study of pre-symptomatic amyotrophic lateral sclerosis (ALS) and the design of disease prevention trials are greatly hampered by our inability to predict which unaffected carriers of ALS-associated pathogenic variants will phenoconvert to clinically manifest disease, and when. In this longitudinal Olink Explore high-throughput proteomic study, 516 serially collected plasma samples from 33 phenoconverters, 35 patients with ALS, 10 pre-symptomatic pathogenic variant carriers and 59 controls were included. We identified 81 proteins whose concentrations changed prior to phenoconversion; characterized the longitudinal trajectory of these proteins; and identified a core panel of 19 proteins that, collectively, predicted phenoconversion over the 0.5- to 5-year time horizons (areas under curve 0.80-0.89) and yielded estimates of time-to-phenoconversion with a mean absolute error of 1.6 years. These findings were replicated in UK Biobank data, confirming pre-symptomatic increases in several proteins (e.g. NEFL, EDA2R, CA3) and that a multi-protein panel outperformed NEFL alone in estimating time-to-phenoconversion. This work sheds light on the biology of pre-symptomatic ALS. Moreover, our identification of a panel of novel susceptibility/risk biomarkers based on empirical longitudinal data furthers the ultimate goal of ALS prevention.

## INTRODUCTION

Historically, neurodegenerative diseases such as amyotrophic lateral sclerosis (ALS), frontotemporal dementia (FTD), Parkinson’s disease (PD) and Alzheimer’s disease (AD) have been defined based on the presence of a characteristic clinical phenotype. Increasingly, however, these disorders are being defined based on their underlying biology ^1–3^, with the recognition that there are both pre-symptomatic and clinically manifest stages of disease. Indeed, there is a growing recognition that early, even pre-symptomatic, intervention is likely to offer the greatest prospect for meaningful therapeutic effect. As a result, a seismic shift – from a traditional care paradigm to prevention – is underway ^4–10^.

While preventing the onset of biologically-defined *disease* might be an aspirational goal, in ALS (and ALS/FTD), emerging efforts are currently focused on preventing *phenoconversion* to clinical manifestations ^7^. A major obstacle to such prevention trials is our limited ability to predict *when* phenoconversion is likely to occur and in *whom* ^8^. The identification of a pre-symptomatic increase in blood neurofilament light chain (NfL) concentration ^11, 12^ was essential for the design and implementation of ATLAS, the first-ever ALS prevention trial ^13^. In the context of this trial, NfL serves as a susceptibility/risk biomarker, predicting risk of phenoconversion at the individual level ^14, 15^. Importantly, the observed rise in NfL in the 6-12 months prior to phenoconversion, was apparent in carriers of highly penetrant *SOD1* pathogenic variants associated with rapidly progressive disease. NfL, however, reflects the rate of axonal loss ^16^, and thus NfL alone may be insufficient for the prediction of phenoconversion among carriers of a broader array of pathogenic variants, especially those that are more slowly progressive ^12, 17^. Also, since NfL reflects axonal loss, a consequence of upstream pathobiological mechanisms of disease, biomarkers reflecting these mechanisms are to be expected. The discovery and validation of additional susceptibility/risk biomarkers is, therefore, critical to the feasibility and success of future disease prevention trials.

Biofluid-based biomarkers in ALS (and ALS/FTD) hold great promise for a host of reasons. First, unlike imaging or physiological-based biomarkers, which largely capture downstream manifestation of disease, biochemical biomarkers have the potential to elucidate the underlying pathobiology of disease. Second, since biofluid-based biomarkers reflect systemic, as opposed to regional or topographically-restricted manifestations of disease, they offer a window into early pathobiological changes irrespective of the anatomical location of disease. Third, there is broad agreement on, and widespread implementation of, standard operating procedures for the collection, processing and storage of biological samples for biomarker development. Finally, emerging omics platforms that have been subjected to rigorous analytic validation permit large-scale and relatively unbiased discovery efforts. Crucial to the endeavor of developing susceptibility/risk biomarkers is the availability of a cohort of individuals (i.e. phenoconverters) who have been prospectively followed over an extended period of time—from the pre-symptomatic state through phenoconversion to clinically manifest disease—and in whom the timing of symptom onset is known and clearly defined. Practically, carriers of pathogenic variants associated with significantly elevated risk for ALS/FTD are currently the only population in whom such natural history studies are feasible.

Over the course of the past ∼18 years, the *Pre-Symptomatic Familial ALS (Pre-fALS)* study ^18^ has followed a large cohort of unaffected pathogenic variant carriers at significantly elevated risk for ALS (or ALS/FTD), with longitudinal data and sample acquisition both before and after phenoconversion to clinically manifest ALS/FTD. Combined with data and biospecimens similarly collected from controls and those with clinically manifest ALS in companion studies, the *Pre-fALS* cohort offers an unprecedented opportunity for the discovery of susceptibility/risk biomarkers to aid the prediction of phenoconversion. Here we report findings from the application of the Olink Explore HT platform to plasma samples from this unique cohort, with independent replication using data from the UK Biobank ^19^.

## RESULTS

### Discovery cohort

This proteomic study included participants from 3 parent studies: *Pre-fALS*, Clinical Research in ALS (CRiALS) Biomarker study, and CReATe Consortium Phenotype-Genotype-Biomarker (PGB1) study. Details of *Pre-fALS* (NCT00317616) have previously been described ^11, 12, 18, 20, 21^. Briefly, *Pre-fALS* is a single-center study that recruits, from across North America, individuals who are carriers of any ALS- (or ALS/FTD)-associated pathogenic variant and who, at the time of enrollment, are clinically pre-symptomatic. CRiALS Biomarker is a companion study to *Pre-fALS*, serving to recruit both healthy controls and patients with clinically manifest ALS, to aid the interpretation of pre-symptomatic data from *Pre-fALS*. CRiALS Biomarker study procedures and clinical assessments mirror those used in *Pre-fALS*. Additional participants with clinically manifest ALS were drawn from CReATe PGB1 (NCT02327845), a multi-center natural history study of individuals with ALS and related disorders, in which participants were evaluated serially to acquire longitudinal phenotypic data and biospecimens. PGB1 participants included in this report were all from the University of Miami site. See Methods for additional details.

### Participant characteristics

For the discovery cohort, 516 plasma samples from N=137 participants were included. The phenoconverter group comprised N=33 *Pre-fALS* participants who had phenoconverted to clinically manifest ALS and/or FTD, and from whom plasma samples and accompanying phenotypic data were available before (N=33) and after (N=23) phenoconversion (total 202 visits). To provide context and facilitate interpretation of data, we also included samples and data from N=59 controls (126 visits), N=35 patients with ALS (126 visits), and N=10 pre-symptomatic pathogenic variant carriers who had not yet phenoconverted (62 visits) (Supplementary Figure S1). With the exception of N=21 controls, all participants contributed longitudinal samples from multiple visits. Demographic and clinical characteristics are summarized in Table 1.

**Table 1.**
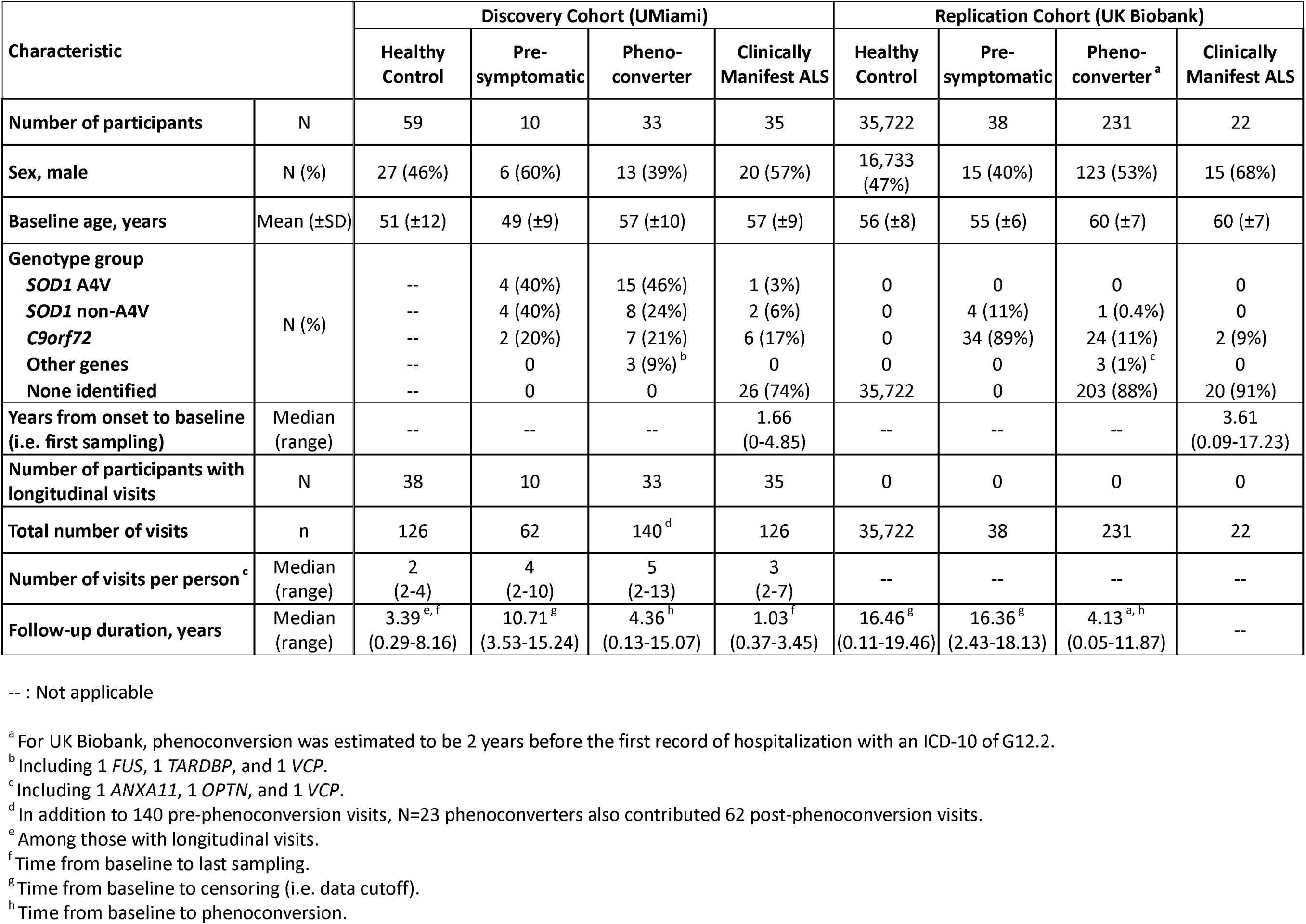
Demographic and clinical characteristics.

### Disease state biomarkers and implicated biological pathways

The Olink Explore HT panel includes 5,440 proteins. After multiple QC steps, data from 5,298 proteins were retained. Applying a mixed-effects model to compare clinically manifest ALS with healthy controls, we identified 137 differentially expressed proteins, of which 105 were upregulated and 32 were downregulated (false discovery rate [FDR] = 0.05) (Figure 1A, Supplementary Table S2). Gene ontology (GO) term ^22^ enrichment analysis identified skeletal muscle as the dominant pathway implicated by upregulated proteins (Figure 1B), and extracellular matrix (ECM) as the dominant pathway implicated by downregulated proteins (Figure 1C). Protein-protein interaction network analysis ^23^ identified clusters related to skeletal muscle function, the ECM, positive regulation of tumor necrosis factor (TNF)-mediated signaling, and regeneration (which includes neurofilament) (Figure 1D). A heatmap depiction of expression patterns of a subset (49 out of the 137) differentially regulated proteins illustrates the distinct molecular signatures associated with disease status (Figure 1E). Parenthetically, in this report we use the non-italicized gene name to refer to the protein product. We adopted this convention for ease of visualization, and to enable comparability to the published literature.

**Figure 1.**
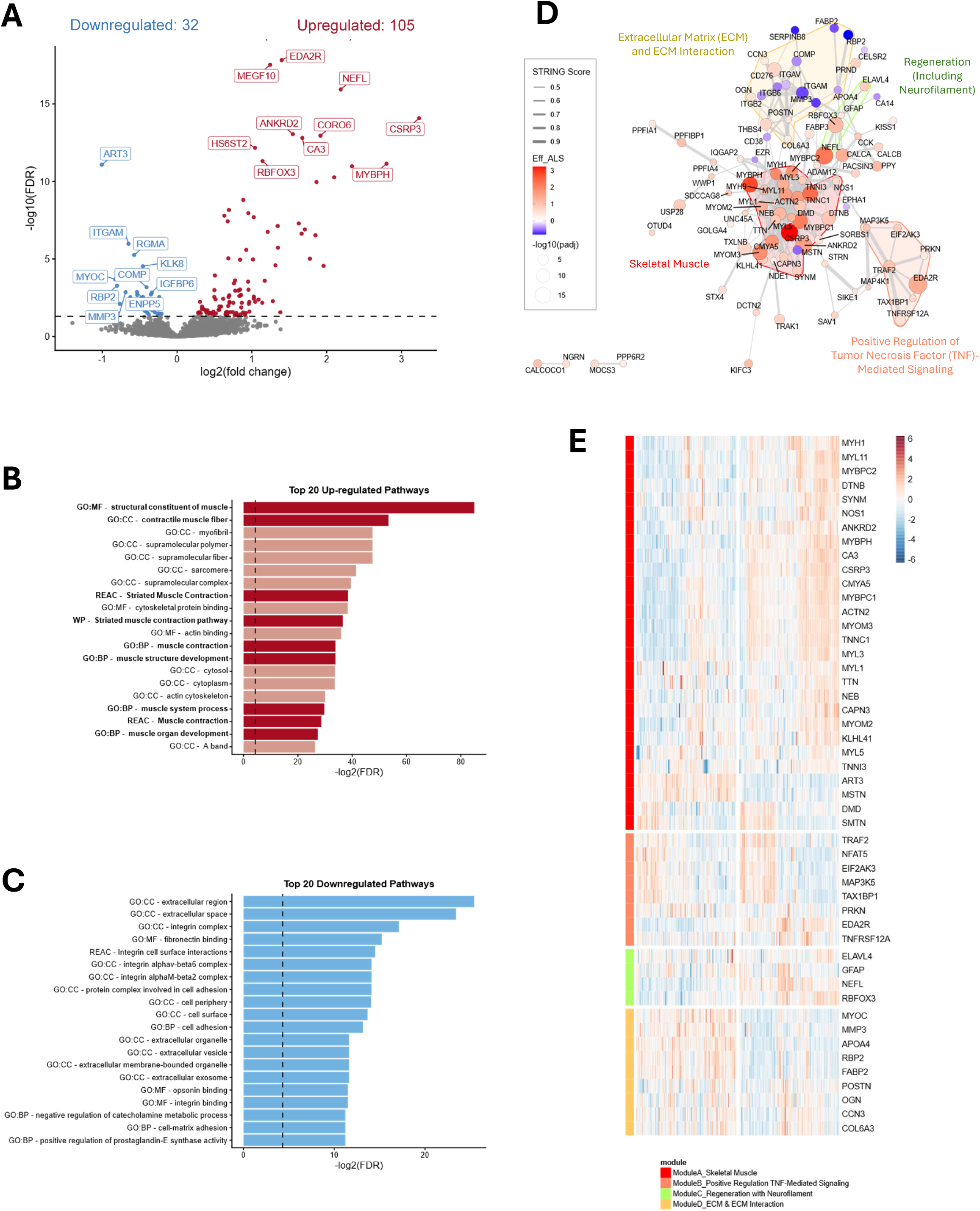
Plasma markers differentially regulated in ALS vs. healthy controls. **(A)** Volcano plot illustrating differentially regulated proteins between clinically manifest ALS and healthy controls. Significantly upregulated proteins are shown in red, and significantly downregulated proteins are shown in blue. The dashed black line indicates the false discovery rate (FDR) threshold of 0.05. **(B)** The top 20 gene ontology (GO) terms that are most enriched among upregulated proteins. Dark red shading and bold font highlights GO terms associated with muscle function. The dashed black line indicates the FDR threshold of 0.05. **(C)** The top 20 GO terms that are most enriched among downregulated proteins. The dashed black line indicates the FDR threshold of 0.05. **(D)** Protein-protein interaction network of differentially regulated proteins, with clusters related to skeletal muscle (red), the extracellular matrix (olive), regeneration and neurofilament (green), and positive regulation of tumor necrosis factor (TNF)-mediated signaling (orange) highlighted. **(E)** Heatmap of the subset of 49 proteins with distinct expression patterns corresponding to the four clusters highlighted in (D). Columns represent data from individual study participants; rows indicate specific proteins.

### Temporal course of biomarkers across pre-symptomatic and clinically manifest ALS

The longitudinal trajectory of biomarkers prior to and following phenoconversion to clinically manifest ALS was examined using data from the phenoconverter and clinically manifest groups. Of the 137 differentially regulated proteins, 81 showed significant changes in their relative abundance (compared to age- and sex-matched controls) before phenoconversion (Figure 2 and Supplementary Figure S2A). Similar trajectory analyses using data from phenoconverters only (including visits before and after phenoconversion) yielded 74 proteins with significant pre-symptomatic change (Supplementary Figure S2B). The same analysis using only phenoconverter data from before phenoconversion yielded 57 proteins with significant change (Supplementary Figure S2C). Procedures for stepwise selection of proteins are summarized in Supplementary Figure S3.

**Figure 2.**
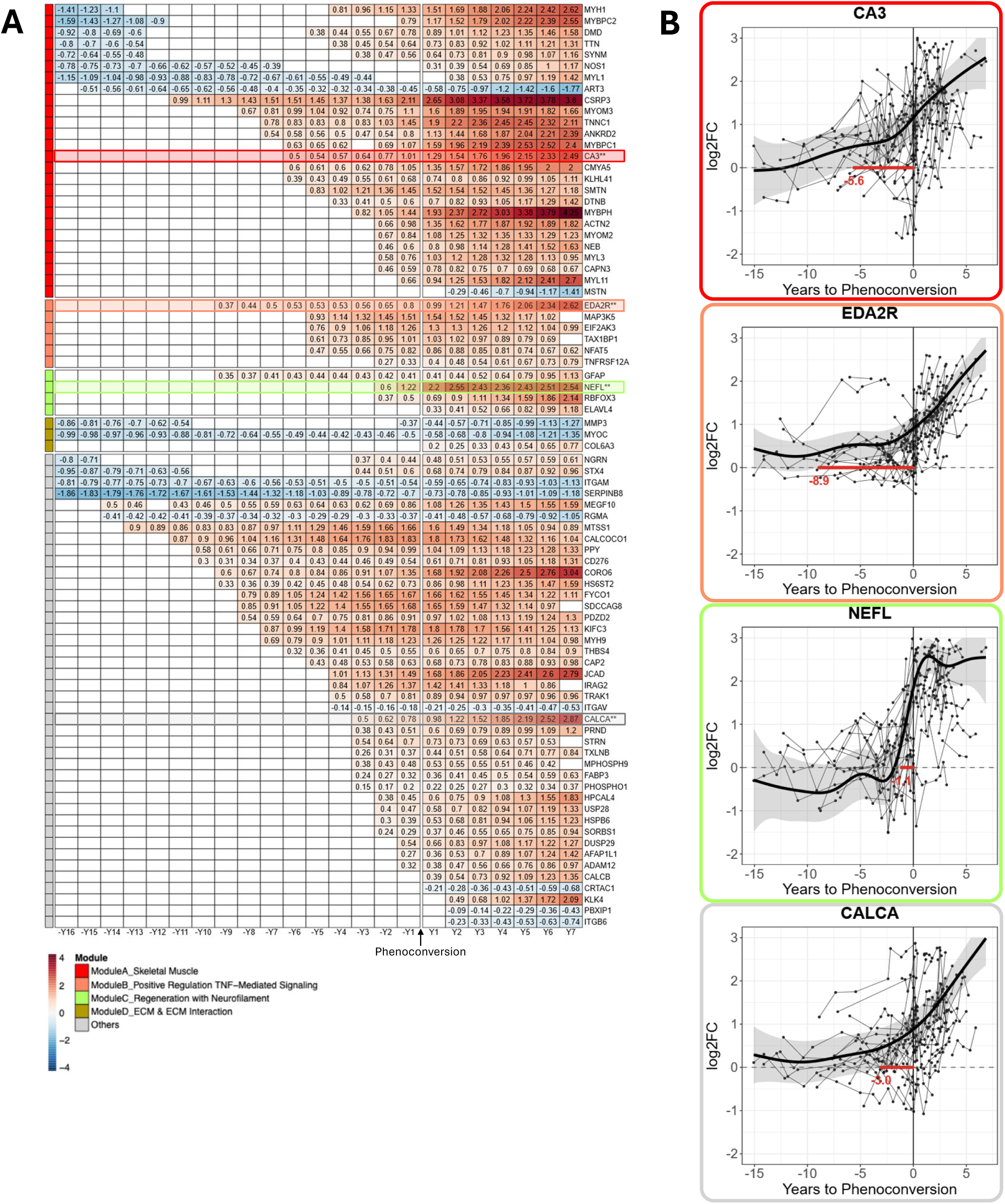
Longitudinal trajectory of protein biomarkers before and after phenoconversion. **(A)** Heatmap depicting 81 proteins with significant changes in their relative abundance, as compared to controls, during the pre-symptomatic and clinically manifest stages of disease. The color scale reflects the direction and magnitude of change, with warmer colors (red) indicating increased concentrations and cooler colors (blue) indicating decreased concentration relative to controls. The number in each cell represents the average protein abundance, relative to controls, during the indicated time interval before or after phenoconversion. Empty cells denote time points where differences were not statistically significant. Proteins are grouped by functional modules (see Figure 1D), as indicated by the color bar in the first column. **(B)** Longitudinal trajectories of four proteins (NEFL, CALCA, EDA2R and CA3), one representing each functional module, to further illustrate relative protein abundance, in log2 fold change (log2FC), during the periods before and after phenoconversion. In addition to phenoconverters, participants with clinically manifest ALS were included; their estimated date of symptom onset was used as a proxy for date of phenoconversion. The dots and connecting lines show the longitudinal data from individual participants. The black curve depicts the estimated mean longitudinal trajectory of protein abundance, adjusting for age and sex; the shaded band denotes the 95% confidence interval around the fitted mean trend. Horizontal red lines indicate the time periods during which relative protein abundance is significantly elevated (i.e. log2FC>0).

Results confirmed prior observations that NEFL levels sharply rise shortly before phenoconversion and continues to increase, albeit more gradually thereafter. In addition, we observed that the concentration of numerous other proteins (e.g. CA3, EDA2R) increased many years before phenoconversion and with further increases during the clinically manifest stage of disease. For a much smaller number of proteins (e.g. ART3, ITGAV), the concentration begins to decline prior to phenoconversion and with further reduction following symptom onset.

### Phenoconversion event prediction

With the goal of building models that accurately predict whether a pre-symptomatic pathogenic variant carrier will phenoconvert to ALS within a certain timeframe, we evaluated the performance of seven machine learning-based classification methods using data from the 137 differentially regulated proteins. We restricted the analysis to these 137 proteins in order to limit the search space given the modest number of phenoconverters. We included proteomic data from the pre-symptomatic group and the phenoconverter group (pre-conversion visits only), and considered 5 different time horizons—that is, whether the participant phenoconverted within 0.5, 1, 2, 3 or 5 years after any given sample collection— with annotation of visits as “cases” or “controls” as described in Methods. The number of “cases” and “controls” and their group affiliation are summarized in Supplementary Table S1. This approach leveraged the unique longitudinal structure of our data; rather than simply predicting whether an individual participant will or will not phenoconvert, this study design enabled us to assess the likelihood of phenoconversion within a specified timeframe given the proteomic profile at each visit. By considering visit (instead of person) level data, we were able to maximize the sample size of our training data to develop better classification models. Optimal 5-fold cross-validation AUCs derived from the seven machine learning algorithms are summarized in Supplementary Figure S4.

We found that logistic regression (LR) models achieved performance comparable to the othertop-performing models, while requiring only a relatively small number of proteins. Using stepwise LR, we then identified, for each timeframe, the model with highest AUC of the receiver operating characteristic (ROC) curve. All subsequent results reported in this section are derived from the LR method with 5-fold cross-validation. The peak predictive performance for each timeframe was: for 6-month prediction, 20 proteins, AUC=0.945; for 1-year, 11 proteins, AUC=0.963; for 2-year, 24 proteins, AUC=0.939; for 3-year, 34 proteins, AUC=0.903; and for 5-year, 25 proteins, AUC=0.897. By comparison, the respective AUC for the model that includes NEFL alone ranged from 0.67 to 0.81 across the five timeframes (Figure 3C). NEFL, not surprisingly, consistently appeared as the strongest single predictive marker across all five timeframes. Interestingly, two other proteins, dual specificity phosphatase 29 (DUSP29) and calcitonin related polypeptide alpha (CALCA), also appeared in the models across five and four timeframes, respectively. The 77 unique proteins that were identified in these models and their overlap across the five timeframes, as well as the respective AUCs, are summarized in Supplementary Table S3.

**Figure 3.**
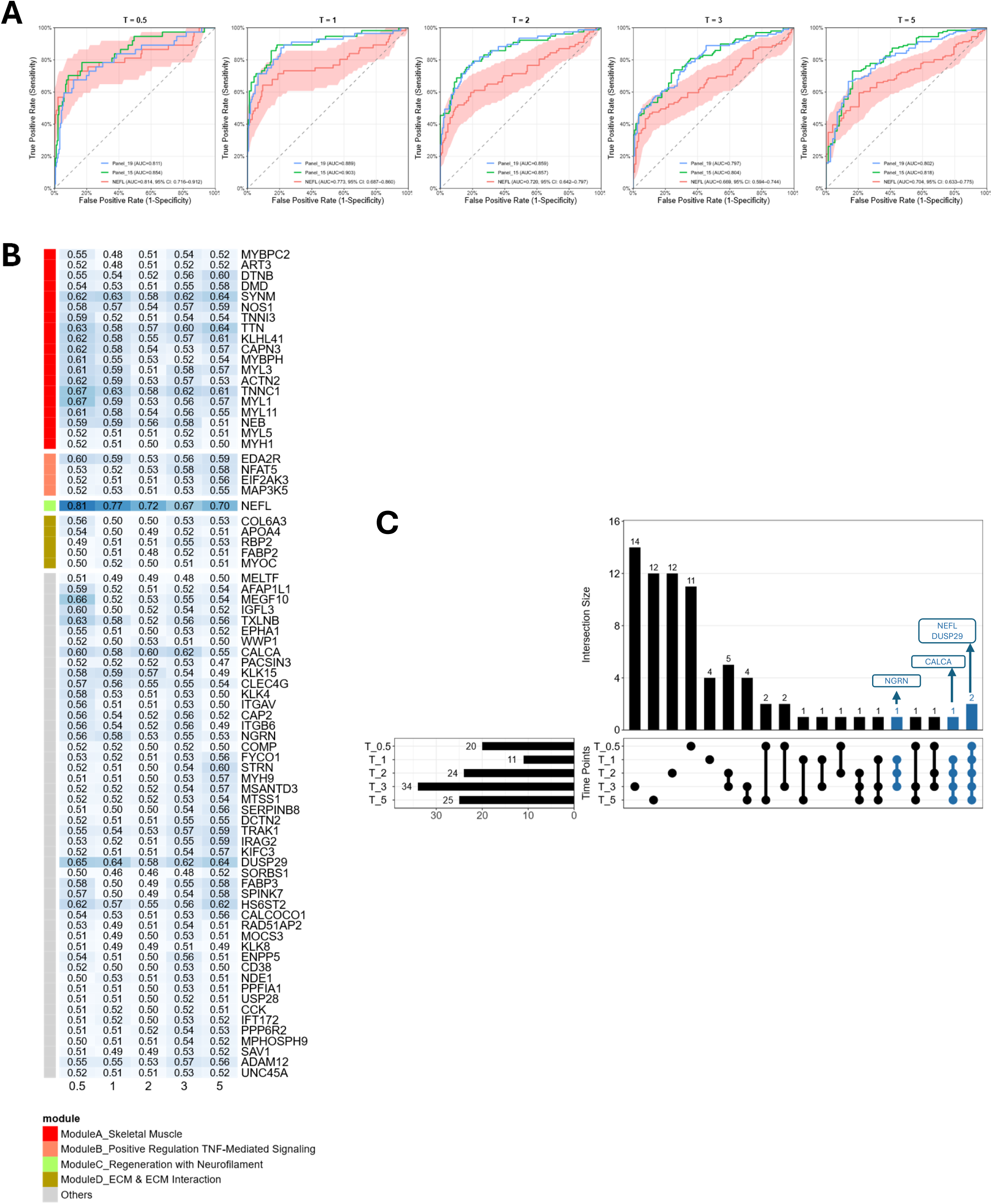
Phenoconversion event prediction. **(A)** Receiver operating characteristic (ROC) curves comparing the performance of phenoconversion event prediction of 3 different models. Red lines correspond to prediction results obtained using NEFL alone (shading represents the 95% CI). Blue lines correspond to results obtained using the 19-protein panel. Green lines correspond to results obtained using the 15-protein panel (subset of 19 proteins that is also available in UK Biobank data). **(B)** Heatmap illustrating the area under the curve (AUC) of ROC obtained, via logistic regression, for each of the 77 proteins. Analyses are conducted across five different timeframes (0.5, 1, 2, 3 and 5 years). Each AUC value is obtained from 5-fold cross validation. Darker shades of blue indicate higher AUC. Proteins are grouped by functional modules (see Figure 1D), as indicated by the color bar in the first column. **(C)** UpSet plot of protein sets identified by logistic regression to predict phenoconversion over the five timeframes. Horizontal bar chart (lower left panel) summarizes the number of proteins included in the model for each timeframe, with NEFL and DUSP29 contributing to phenoconversion event prediction across all 5 timeframes.

From these 77 proteins we selected a subset to form a core panel of proteins, using a data-driven approach supplemented by expert curation. We considered the ranking of each protein for phenoconversion prediction within each time interval, favoring those ranked more highly, and sought to ensure a balance of biomarkers between those that were predictive of phenoconversion across time periods and those only predictive for a single time period (Supplementary Table S3). This led to the compilation of a 19-protein biomarker panel that included: NEFL, DUSP29, CALCA, NEB, NGRN, MYL11, NOS1, MYH1, TNNC1, DTNB, MEGF10, SYNM, APOA4, ART3, MYL3, EPHA1, EDA2R, TTN, and ACTN2.

In predicting phenoconversion, this 19-protein panel (Figure 3A) outperformed that of any single protein for any timeframe (Figure 3B), underscoring the need for a multi-biomarker approach to forecasting phenoconversion across both short- and intermediate-term timeframes. There was, however, one exception: for predicting phenoconversion within 0.5 year, the 19-protein panel (which included also NEFL) and NEFL alone performed comparably, suggesting that the dramatic rise in NEFL in the months immediately before phenoconversion may be sufficient for short-term prediction (Figures 3A). The specific protein sets selected for each timeframe and their overlap across models are summarized in Supplementary Table S3 and illustrated in Figure 3C, revealing shared and distinct biomarkers associated with different time periods prior to phenoconversion. In addition, for comparison, Figure 3A also shows the AUC of a 15-protein subset of these 19 proteins, selected based on their availability in the UK Biobank data, which we used for replication analysis (see below and Supplementary Table S3).

### Phenoconversion timing estimation

In addition to predicting binary phenoconversion status within specified timeframes, we applied linear mixed effects models with truncation regression to estimate time-to-phenoconversion among phenoconverters. Age, sex and genotype group were included as covariates. For the 19-protein panel, the correlation (cor) between estimated and actual years-to-phenoconversion was 0.79, with a mean absolute error (MAE) of 1.62 years (Figure 4A). A 15-protein panel (to facilitate replication analysis using UK Biobank data – see below) yielded similar but slightly inferior results (Figures 4B, cor=0.76, MAE=1.72). Compared to NEFL alone (Figures 4C, cor=0.48, MAE=2.37), however, the 19-protein panel performed meaningfully better.

**Figure 4.**
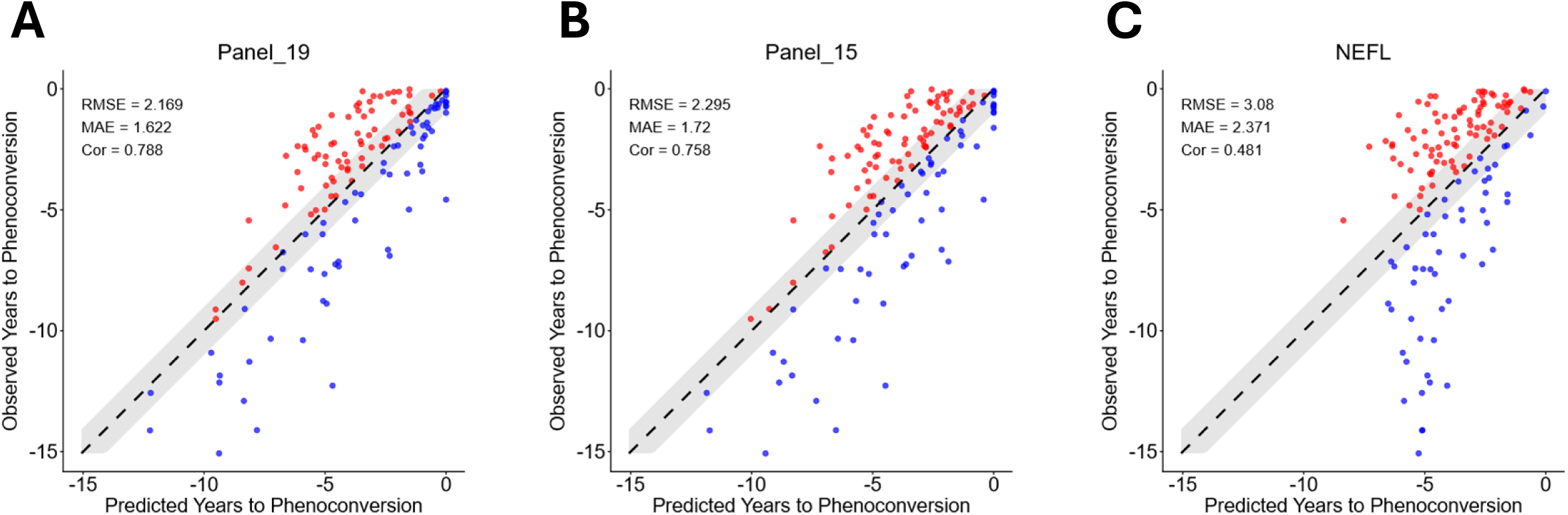
Time-to-phenoconversion estimation. Scatterplots illustrating the relationship between observed and estimated (or predicted) time-to-phenoconversion for **(A)** the 19-protein panel, **(B)** the 15-protein panel (subset of 19 proteins that is also available in UK Biobank data), and *(C)* NEFL alone. Red dots indicate overestimation and blue dots indicate underestimation. RMSE = root mean square error (average error between predicted and observed); MAE = mean absolute error (average absolute difference between predicted and observed); Cor = Pearson correlation coefficient.

### UK Biobank replication

For replication analysis, we utilized data from the UK Biobank Pharma Proteomics Project (UKB-PPP). Using inclusion/exclusion criteria that yielded participants comparable to the four groups in the discovery cohort, we identified from UKB-PPP N=35,722 healthy controls, N=38 pre-symptomatic *SOD1* and *C9orf72* carriers, N=231 phenoconverters, and N=22 clinically manifest ALS for inclusion in the replication cohort (Table 1, Supplementary Figure S5; see Methods for additional details). The top disease state biomarkers identified in the discovery cohort largely overlapped with those identified in the replication cohort (Figure 5A). Notwithstanding some key differences between the two cohorts – proteomic data from UK Biobank were cross-sectional and included ∼3000 proteins (Olink Explore 3072), whereas our data were longitudinal and included >5400 proteins (Olink Explore HT) – comparing phenoconverters to age- and sex-match controls, we observed in the replication cohort similar temporal patterns as we did in the discovery cohort for a subset of proteins (NEFL, EDA2R, TNFRSF12A, CA3, DTNB, HSPB6, and ITGB6) (Figures 5B). Notably, 15 of the 19 core proteins identified in the discovery cohort were included in Olink Explore 3072, constituting a 15-protein panel. The trajectories of NEFL and EDA2R are illustrated in Figure 5C. Importantly, these trajectories in the replication cohort are only pseudo-longitudinal, as they are compiled from cross-sectional data. Using the 15 proteins to fit a truncated regression model to estimate years to phenoconversion, we found that the 15-protein panel outperformed NEFL alone, with MAEs of 2.75 and 3.61 years, respectively (Figure 5D).

**Figure 5.**
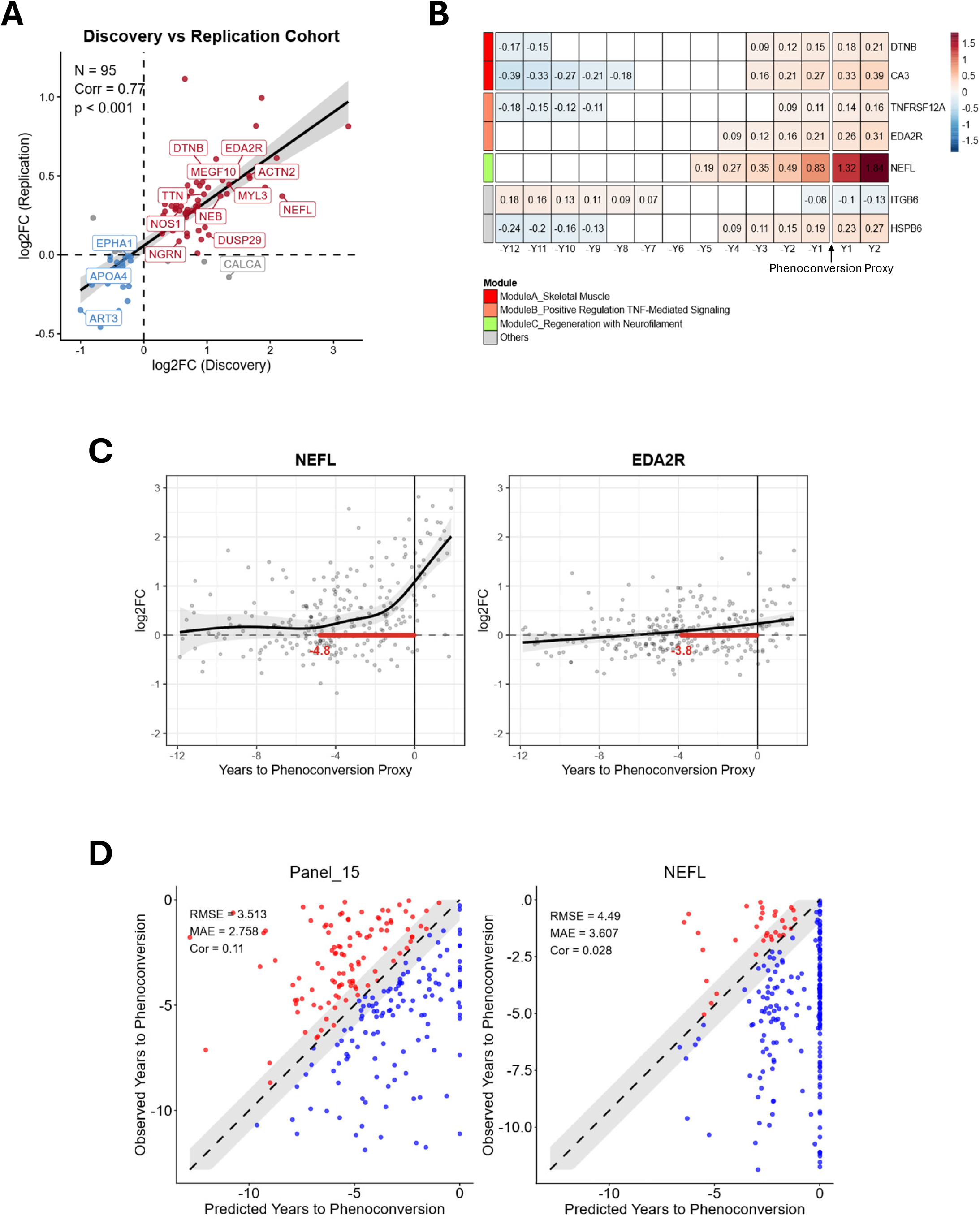
UK Biobank replication. **(A)** Scatterplot showing high correlation between discovery and UKB replication cohorts, for proteins that were differentially (up and down) regulated in clinically manifest ALS vs. healthy controls. **(B)** Pseudo-longitudinal (constructed using cross-sectional data) patterns of 7 proteins with relative abundance that deviates significantly from healthy controls. This heatmap included data from phenoconverters and those with clinically manifest ALS. The number in each cell represents the average relative protein abundance at the indicated time interval, with negative and positive time being, respectively, before and after phenoconversion. Empty cells denote time intervals where differences were not statistically significant. The color scale reflects the direction and magnitude of change, with warmer colors (red) indicating increased concentrations relative to controls and cooler colors (blue) indicating decreased concentration relative to controls. Proteins are grouped by functional modules (see Figure 1D), as indicated by the color bar in the first column. **(C)** Visual depiction of pseudo-longitudinal trajectories of two proteins of interest (NEFL and EDA2R) relative to sex- and age-matched controls. Horizontal red bars indicate the time periods during which the relative protein abundance is significantly elevated (i.e. log2FC > 0). The grey shaded areas represent the 95% confidence interval for the relative protein abundance. **(D)** Scatterplots illustrating the relationship between observed and estimated time-to-phenoconversion for the 15-protein panel and for NEFL alone in UKB data.

## DISSCUSSION

This study is the first to map the longitudinal trajectory of an array of protein biomarkers across the course of ALS disease progression, from pre-symptomatic through clinically manifest stages of disease. Prior studies have relied on cross-sectional proteomic measures ^24, 25^, have made assumptions about the timing of phenoconversion ^24, 25^, or have inferred pre-symptomatic biomarker trajectories based on data from pre-symptomatic and clinically manifest individuals rather than phenoconverters ^17^. By contrast, our findings derive from longitudinal proteomic data from the largest cohort of phenoconverters evaluated both prior to and following the emergence of clinically manifest disease, and with the timing of phenoconversion rigorously determined. In univariate analyses, we found that the concentration of 81 proteins deviated from age- and sex-matched controls prior to phenoconversion, with skeletal muscle, the extracellular matrix, neurofilament and regeneration, as well as positive regulation of TNF signaling among the biological pathways most impacted. In addition to shedding light on the biology of pre-symptomatic disease, we have identified a 19-protein panel that predicts the event of phenoconversion with high accuracy (AUC 0.80-0.89) across a range of time horizons from 0.5 to 5 years. Critically, these AUCs are not artificially inflated by comparing phenoconverters to healthy controls – but rather, they reflect the real-world challenge of differentiating phenoconverters from pre-symptomatic carriers who do not phenoconvert within the specified timeframe. These findings build on our prior observation of NfL’s utility as a susceptibility/risk biomarker predicting phenoconversion to clinically manifest ALS. Importantly, while NfL alone may be sufficient over the short-term among carriers of highly penetrant *SOD1* variants associated with rapidly progressive disease, additional susceptibility/risk biomarkers are needed for longer range prediction and for those with less aggressive forms of disease. Moreover, we have made significant inroads into the challenge of estimating the years-to-phenoconversion among pathogenic variant carriers which, until now, has been impossible to predict. Using linear mixed effects models and our 19-protein panel (which includes NEFL) data from phenoconverters, the only group in whom the timing of phenoconversion is definitively known, we are able to estimate years-to-phenoconversion with a mean absolute error (MAE) of 1.6 years. The ability to estimate time-to-phenoconversion should further empower the study of pre-symptomatic ALS by providing a temporal anchor for interpretating data from carriers who have not phenoconverted.

In constructing a multi-protein panel to serve as a susceptibility/risk biomarker, we considered the pros and cons of adopting a purely data-driven approach vs. a supervised data-driven approach augmented by expert curation. While the former is unbiased and is more likely to maximize predictive performance, this approach is also susceptible to overfitting, poor reproducibility, and limited biological interpretability. On the other hand, the latter (which is the approach we have taken) risks reinforcing established ideas and may have lower predictive power. However, it benefits from yielding a result that is biologically more plausible, interpretable, and with better reproducibility ^26^.

We recognize the intrinsic bias of the targeted Olink platform, in which well known, higher abundance, extracellular proteins are overrepresented. Nevertheless, our results are consistent with prior studies in identifying NEFL as one of the most differentially regulated proteins when comparing those with clinically manifest ALS to healthy controls. Of all the proteins examined, NEFL is also the single most useful biofluid-based biomarker of pre-symptomatic disease. However, the largest number of proteins identified as differentially regulated between ALS and controls, as well as those that change pre-symptomatically and contribute to predicting the timing of phenoconversion, are not neuronal; rather, they reflect the biology of skeletal muscle. Specifically, many of these are structural components of skeletal muscle: MYH1 and MY11 (highly expressed in fast-twitch fibers); MYL3, CRSP3, ANKRD2, and TNNC1 (slow-twitch); as well as TTN, NEB, SYNM, and ACTN2 (sarcomeric). Several, on the other hand, reflect cellular processes related to skeletal muscle atrophy (DUSP29, EDA2R, MEGF10, CALCA). DUSP29 is noteworthy as it is transcriptionally activated in denervation-induced atrophy, attenuating the ERK1/2 branch of the MAP kinase signaling pathway and inhibiting muscle cell differentiation ^27^; DUSP29 was also identified as a pre-symptomatic marker based on SomaScan data in the EPIC cohort ^25^. Similarly, EDA2R, a member of the TNF receptor superfamily, has emerged as a significant mediator of skeletal muscle atrophy ^28^. A loss of NOS1, which inhibits FOXO3, a transcription factor that induces proteolytic and autophagic pathways, is an early event in skeletal muscle atrophy ^29^. MEGF10 is essential for skeletal muscle development and maintenance, with deficiency known to result in loss of muscle mass ^30^. Similarly, CALCA, which is released from motor neurons and has a role in inhibiting autophagic lysosomal proteolysis, likely reflects an anabolic response to denervation-induced atrophy. While the observed increase in MEGF10 and CALCA likely reflect a compensatory response to neurogenic atrophy, the increase in EDA2R and DUSP29 is unexpected and may reflect a role in driving or exacerbating muscle atrophy. The observation that CALCA is elevated in ALS vs. controls and among pathogenic variant carriers prior to ALS phenoconversion is also of interest given studies that have identified increased expression of calcitonin gene related peptide as a marker of motor neuron vulnerability in the SOD1^G93A^ mouse ^31^.

The pre-symptomatic increase in EAD2R, MAP3K5, EIF2AK3, TAX1BP1, NFAT5 and TNFRSF12A suggest increased inflammatory cytokine signaling in response to oxidative and endoplasmic reticulum (ER) stress, with activation of both pro-apoptotic and cell survival pathways. This observation is consistent with what is understood about early events in the pathogenesis of ALS, where the accumulation of misfolded proteins leads to ER stress, which in turn leads to activation of the unfolded protein response (UPR) ^32^. Here we provide the first *in vivo* human data supporting early activation of these cellular processes, temporally preceding the rise in NfL.

Major strengths of this study include the use of longitudinal proteomic data and detailed information about the timing of phenoconversion, in the largest available series of phenoconverters with such data available. A limitation of our study, however, is the lack of an independent, but comparable, cohort for replication. Nevertheless, we were able to utilize Olink data from the UK Biobank study, which includes whole genome sequencing and proteomic data from participants who subsequently developed (mostly sporadic) ALS. A key challenge was that the exact timing of phenoconversion is not available in UK Biobank data. Instead, we relied on ICD-10 codes for motor neuron disease (MND, G12.2) at the time of hospitalization, as well as death records with ALS recorded as contributing to death. Since neither hospitalization nor death are early events in the course of ALS, these data do not readily lend themselves to replication of an analyses focused on phenoconversion. To partially mitigate this issue, we assumed onset to be 2 years before hospitalization with an associated ICD-10 code of G12.2 and use this as the proxy timepoint of phenoconversion. Despite the imperfections of this approach, we were able to replicate the findings of NEFL, EDA2R, CA3, TNFRSF12A, DTNB, ITGB6 and HSPB6 as pre-symptomatic biomarkers, and showed that a multi-protein panel improved estimates of the predicted time-to-phenoconversion above and beyond what can be accomplished using NEFL alone. That the results from our discovery cohort were only partially replicated in the UK Biobank replication cohort, is likely due in part to the limitations of relying on cross-sectional proteomic data and very imprecise proxies of the timing of phenoconversion.

In modeling the pre-symptomatic trajectory of protein biomarkers, we have utilized data from phenoconverters with longitudinal measurements both before and after phenoconversion, as well as data from those with clinically manifest ALS. It is noteworthy that estimates of pre-symptomatic trajectories differ somewhat when, instead, relying solely on longitudinal data from phenoconverters pre- and post-conversion, or when using only pre-conversion data from phenoconverters. Moreover, models that rely solely on data from pre-symptomatic carriers who have not yet phenoconverted (and in whom the timing of phenoconversion is unknown) and data from those with clinically manifest ALS (in whom the timing of phenoconversion is retrospectively estimated) are inherently less robust.

As other ongoing pre-symptomatic studies accrue larger number of phenoconverters and more longitudinal visits, and large-scale proteomic analyses become more accessible, it will be important to more fully replicate our findings. Moreover, practical utilization of a panel of proteins as a susceptibility/risk biomarker will benefit from immunoassay validation of leading candidates, as well as demonstration that the results of such immunoassays hold sufficient predictive value for use as eligibility criteria for enrollment or randomization. Nevertheless, the results reported here are immediately relevant to the design of future ALS prevention studies. Specifically, clinical trials that aim to prevent phenoconversion to clinically manifest ALS/FTD will need to enrich the study population for those at greatest short-term risk of the primary outcome measure that will be used to quantify treatment effect. As such, our identification of a panel of proteins that reliably predict the timing (and occurrence) of phenoconversion, represent an important advance in furthering the ultimate goal of ALS prevention.

## METHODS

Overall study design and statistical methods are summarized in Supplementary Figure 1. The discovery cohort comprised participants from the parent studies at the University of Miami (described below). The replication cohort was drawn from the UK Biobank. As previously described, we regard the pre-symptomatic stage of disease to be from the onset of the underlying disease (biologically-defined) to phenoconversion ^8, 9, 33, 34^. Throughout this manuscript we use the non-italicized gene name to refer to the protein product. We adopted this convention for ease of visualization, and to enable comparability to the published literature.

### Parent studies: cohort descriptions, ethics approvals, sample collection and processing

This proteomic study included participants from 3 parent studies: *Pre-fALS*, Clinical Research in ALS (CRiALS) Biomarker study, and CReATe Consortium Phenotype-Genotype-Biomarker (PGB1) study. Details of *Pre-fALS* (NCT00317616) have previously been described ^11, 12, 18, 20, 21^. Briefly, *Pre-fALS* is a single-center study that recruits, from across North America, individuals who are carriers of any ALS- (or ALS/FTD)-associated pathogenic variant and who, at the time of enrollment, are clinically pre-symptomatic for ALS and FTD. Since its inception in 2007, *Pre-fALS* has been broadly inclusive regardless of participants’ geolocation and genotype, to maximize the representativeness of the study cohort and generalizability of the results. Participants undergo screening ^18^ as well as genetic testing and counseling ^20^ as needed. Eligible participants are enrolled in the *Pre-fALS* Cohort and followed longitudinally, with detailed phenotypic evaluations, biospecimen collections, and multi-modal biomarker procedures. If evidence of clinically manifest disease is found during baseline evaluation, the individual is excluded from the *Pre-fALS* Cohort and enrolled, instead, in the CRiALS Biomarker study (see below) and referred clinical care. Notably, we do not regard these individuals as phenoconverters, a designation reserved for those who were pre-symptomatic at baseline and subsequently developed clinically manifest ALS or FTD. Among Cohort enrollees, the subset who choose not to learn their genetic test results (“non-disclosure” group) ^18^ comprise those who do and do not carry a pathogenic variant; the latter subgroup are effectively controls. CRiALS Biomarker is a companion study to *Pre-fALS*, serving to recruit both healthy controls and patients with clinically manifest ALS, to aid the interpretation of pre-symptomatic data from *Pre-fALS*. CRiALS Biomarker study procedures and clinical assessments mirror those used in *Pre-fALS*. Additional participants with clinically manifest ALS were drawn from CReATe PGB1 (NCT02327845), a multi-center natural history study of individuals with ALS and related disorders, in which participants were evaluated serially to acquire longitudinal phenotypic data and biospecimens. PGB1 participants included in this report were all from the University of Miami site.

All 3 studies were approved by the University of Miami institutional review board (IRB), which also serves as the single IRB of record for the CReATe Consortium, and all study participants provided written informed consent.

Plasma samples were collected, processed, and stored according to strict standard operating procedures. Briefly, blood was collected in K2 EDTA tubes, centrifuged at 1,750 x g for 10 minutes at 4°C, and aliquoted for storage at −80°C.

### Experimental design: participant and sample selection, Olink plate assignment

An experiment of 516 plasma samples was planned, with a focus on phenoconverters, but also including pre-symptomatic pathogenic variant carriers who have not developed ALS as well as both healthy controls and patients with clinically manifest ALS.

All *Pre-fALS phenoconverters* with at least 2 plasma collections at the time of sample selection were included. A small number of *pre-symptomatic* participants thought to be of higher likelihood of phenoconversion in the near future, based on clinical or biomarker data, were also selectively included. The rationale for their selection was the potential to increase the number of phenoconverters by the time of data analysis (notwithstanding that this particular subset of pre-symptomatic carriers might bias some of the results towards the null). Indeed, two of them phenoconverted before final data analyses (data cutoff: January 2025), and were included in this report as phenoconverters. *Controls* with known comorbidities were excluded; and if a control participant had more than 3 plasma collections, the first and last collection along with a collection near the mid-point of study follow-up were included. (One exception: N=1 control had 4 collections included.) The *clinically manifest* group included those with at least 3 plasma collections (exceptions: N=2 each had just 2 samples with sufficient volume available) and were broadly representative of patients at different stages of disease. Plasma collected at time of known or possible exposure to ASOs (through clinical administration or clinical trials) were excluded. The groups were age- and sex-matched as much as possible.

In the Olink experiment, the 516 plasma samples (40µL each) were run on 6 plates of 86 samples each. Longitudinal samples from the same participant were included on the same plate. In addition, plate assignment was devised to achieve a balanced design, with similar distributions of participant group (control, pre-symptomatic, phenoconverter, clinically manifest), age, and sex on each of the 6 plates.

### Proximity extension assay, library preparation and next-generation sequencing

Olink library preparation and sequencing were performed using the Explore proximity extension assay (PEA) technology and the Explore HT protein biomarker panel. PEA was performed as per the proteomic method previously described ^35^. Briefly, the PEA technology employs high-multiplex matched pairs of antibodies, each conjugated to a unique DNA oligonucleotide. These antibodies were incubated with their respective target proteins, facilitating specific binding. Upon hybridization of antibodies, DNA polymerase-mediated extension generated a unique DNA barcode for each antibody-target interaction wherever both specific antibodies recognized the same protein. The resulting DNA barcodes were amplified using polymerase chain reaction (PCR), incorporating unique sample indexes to enable multiplexing of samples. Following amplification, Olink libraries were purified using Agencourt AMPure XP magnetic beads to remove unwanted PCR byproducts. The quality of the purified libraries was assessed using an Agilent TapeStation system to ensure proper fragment size distribution and integrity. Each assay has been extensively validated for limit of detection, measurement ranges, precision, reproducibility, and specificity, as previously described ^36^. The prepared Olink libraries were sequenced using S4 flow cells on the Illumina’s NovaSeq 6000 platform. Data underwent quality control (QC) checks to ensure sequencing accuracy and integrity.

### Normalizing protein expression

Olink normalized protein expression (NPX) represents the relative protein concentration units in log2 scale. The values are calculated from the number of sequencing reads matching the reference Olink protein barcodes. Data is then normalized to internal extension controls, spiked in during the sample preparation and then log2 transformed. As the study involves more than a single plate of samples, the data for the whole cohort was then normalized to remove plate to plate variations. Briefly, for each plate and assay, the plate-specific median value is calculated then the plate-specific median is subtracted from every sample of the plate, this centralizes the median to 0. Illumina raw data was converted to counts using Olink’s NGS2counts pipeline and NPX calculations and QC analysis was performed with Olink’s NPX Explore HT software.

### Statistical analysis

We applied machine learning methods to predict the occurrence and timing of phenoconversion. Our method for discovery progressed through four steps: differentially expressed proteins selection, temporal dynamics protein modeling, phenoconversion event prediction, and phenoconversion timing estimation. All statistical analyses were performed in R 4.4.0.

#### Step 1. Differentially expressed proteins selection

The goal in this step was to select a subset of differentially expressed proteins (i.e. disease state biomarkers) from the >5,400 in the Olink Explore HT panel for downstream analysis. This step was based on the rationale that proteins differentially expressed between clinically manifest ALS patients and healthy controls are likely ALS-related and would be strong candidates to predict phenoconversion among pathogenic variant carriers. To ensure no overlap between samples used in this step and those used to build prediction models, we included in this step only study participants from the clinically manifest (*N* = 35) and healthy control (*N* = 59) groups, each with Olink data from 126 visits (Table 1).

Since all but 21 controls contributed multiple samples, we employed a mixed-effects model with a random intercept to account for within-person correlation. We also included age at sample collection, sex, and genotype group (*SOD1* A4V, *SOD1* non-A4V, *C9orf72*, other pathogenic variants) as covariates. The Benjamini-Hochberg (BH) procedure was applied to control the false discovery rate (FDR) at a significance threshold of 0.05.

#### Step 2. Temporal dynamics protein modeling

To evaluate whether the differential abundance observed between clinically manifest ALS patients and healthy controls can also be detected prior to phenoconversion, we modeled the longitudinal trajectories of the subset of proteins identified in Step 1, using a two-layer generalized additive mixed model (GAMM) framework ^37^.

In the first layer, we modeled the protein trajectory in healthy controls only and included sex as a fixed covariate, a random intercept to account for within-subject correlation, and a smooth function of age 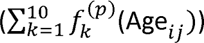 to capture non-linear age-related changes in protein abundance:

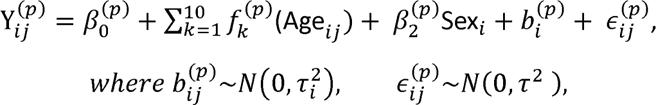

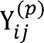 is the observed normalized abundance of protein *p* for individual *i* at visit *j,k* is the number of component functions (range: 1-10), *b* is the individual random effect, and is the random error.

In the second layer, we modeled the relative protein abundance (*δ*) over time, with relative protein abundance defined as the deviation from the expected protein abundance in an age- and sex-matched healthy control based on model in the first layer. The GAMM model in the second layer included sex and genotype group as fixed covariates, a random intercept for individual-level variation, and a smooth function 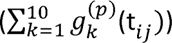 of time to phenoconversion (*t_ij_*). This allowed us to characterize protein trajectory patterns as each individual approached phenoconversion.

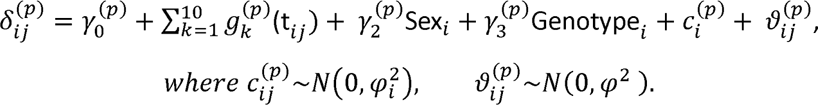

To assess the statistical significance of the disease effect in each model, we applied the BH procedure on smooth term to control the false discovery rate at a significance threshold of 0.05.

These models included data from both phenoconverters and those with clinically manifest ALS (Supplementary Figure S2A). For the latter group, reported onset of weakness was used as a proxy for phenoconversion. Secondarily, we repeated these models but included only data from phenoconverters (pre- and post-conversion) (Supplementary Figure S2B) or only pre-conversion visits from phenoconverters (Supplementary Figure S2C).

#### Step 3. Phenoconversion event prediction

Using the differentially expressed proteins (i.e., disease state biomarkers) identified in Step 1, Step 3 aimed to build prediction models that accurately predict whether a pre-symptomatic pathogenic variant carrier will phenoconvert to ALS within a certain timeframe (T), with T ranging from 0.5 to 5 years. Training data were drawn from the pre-symptomatic and phenoconverter groups, with each visit annotated as either “case” or “control”, depending on, respectively, whether or not phenoconversion occurred within the specified time period following the visit. For the pre-symptomatic group, all visits with subsequent follow-up time longer than (or equal to) T were annotated as “controls”; the remaining visits were excluded as we do not know whether the participant might still phenoconvert within the T year interval. For the phenoconverter group, all visits where phenoconversion occurred more than T years after the visit were considered “controls”, whereas all visits where phenoconversion occurred within T years of the visit were considered “cases”. Each phenoconverter’s first post-conversion visit was also included and labeled as a “case,” while all other post-conversion visits were excluded.

We evaluated seven machine learning algorithms (logistic regression, support vector machine, boosting, random forest, XGBoost, Elastic Net, and LASSO), including age, sex and genotype group as covariates, and found that logistic regression (LR) models achieved high predictive performance using only the relatively small number of proteins from Step 1; the LR models were also more robust given our modest sample size, and their results readily interpretable (Supplementary Figure S4). Model training and testing were performed using five-fold cross-validation. Training and testing sets were stratified to maintain a balance of cases and controls. Additionally, to maintain independence between the training and test sets (i.e. to prevent leakage across folds), we ensured that visits from the same participant did not appear in more than one of the five folds.

We tested five different timeframes: T = 0.5, 1, 2, 3 and 5 years. For each timeframe, we first ran univariate LR for each protein individually, with covariates for sex, age (at sample collection) and genotype group. From the results, we identified the proteins that returned the best prediction measured by area under the curve (AUC) of a receiver operating characteristic (ROC) curve. Next, we performed a greedy stepwise LR with forward selection. Specifically, we progressively added, one at a time, proteins that maximized the incremental (or minimized the decremental) predictive performance among all remaining proteins. Performance was measured using the average AUC across the five-fold cross validation. The model with the best predictive performance in the testing set was selected as the designated model for that timeframe. We counted how many times each of the differentially expressed proteins from Step 1 are included in the five designated models (one for each timeframe), and ranked these proteins based on the number of appearances (Supplementary Table 2) across the 5 models. Among the proteins that appeared at least once in the five models, we further filtered them based on their ranking, the timeframe(s) in which they appeared, as well as biological rationale to arrive at a small core panel of proteins as susceptibility/risk biomarkers.

#### Step 4. Phenoconversion timing estimation

Our goal in this step was to build disease progression models (DPM) to estimate pre-symptomatic mutation carriers’ proximity to time of phenoconversion given the proteomic profile obtained from the individual’s most recent visit and baseline visit, similar to the approach employed in a recent study on familial FTD that utilized a Bayesian DPM ^17^. For this Step, we used the final protein panel identified in Step 3. Instead of LR, we used a truncated regression model and included only data from the pre-conversion visits of phenoconverters, the only group in whom time-to-phenoconversion (*t_ij_*) is known. (Note that *t_ij_* is negative for visits before phenoconversion.) To avoid the multicollinearity between age and time-to-phenoconversion, we used relative protein abundance from Step 2 as input features. Given the longitudinal structure of the proteomics data, with repeated measurements collected from the same individuals over time, we used each participant’s baseline visit as internal reference to account for inter-individual variability in protein abundance. To model time-to-phenoconversion as a continuous outcome, we employed truncated regression with an upper bound at zero, ensuring that all predicted values *t̂_ij_* were negative. In addition, we included sex and genotype group as covariates in the model:

For individual *i* at visit *j*,

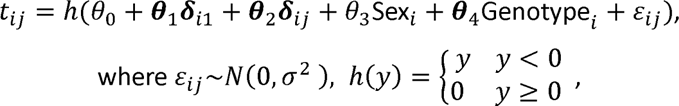

*δ_i_*_1_ is the relative protein abundance vector for individual *i* at baseline visit, and

*δ_ij_* is the relative protein abundance vector for individual *i* at visit *j*.

#### Pathway analysis

Pathway enrichment analysis was performed using the ‘gost’ function in the gprofiler2 package (version 0.2.3), which provides functional profiling by mapping differentially expressed proteins to Gene Ontology (GO), KEGG, Reactome, and other biological databases ^38^. Statistical significance was defined as an FDR-adjusted P value < 0.05.

#### Protein-protein interaction analysis

Protein-protein interaction (PPI) networks were constructed to investigate the functional connectivity among the differentially expressed proteins. Interactions were queried using the STRING database (version 11.5), which integrates evidence from experimental data, computational prediction, co-expression, curated databases, and literature mining ^23^. To identify biologically coherent subnetworks, we applied Markov Cluster Algorithm (MCL) clustering using STRING’s built-in implementation, with an inflation parameter set to 3 to control cluster granularity.

### UK Biobank replication

For replication analysis, we used data from the UK Biobank (UKB) cohort. UKB is a large-scale prospective study that recruited 503,317 participants aged 40-69 years between 2006 and 2010 across the United Kingdom ^19, 39^, with Olink Explore 3072 data available through the UKB Pharma Proteomics Project (UKB-PPP) for N=54,219 ^40^. Data for the present analysis were accessed under application #847687.

The replication cohort consists of four participant groups, which are similarly defined as in the discovery cohort: heathy control, pre-symptomatic, phenoconverter, and clinically manifest (Table 1). From among the UKB-PPP participants, we included only the N=52,488 with also whole genome sequencing data available and who had neither withdrawn consent nor been lost to follow-up (field 190) as of time of data access in August 2025 (Supplementary Figure S5).

Among them, N=327 ALS cases were identified using the International Classification of Diseases (ICD-10) code of G12.2 (motor neuron disease; field 41270) based on self-report at the time of enrollment, or in subsequent hospitalization or death records ^41^. From this group, we included only those whose G12.2 codes were from hospitalization records with exact dates (N=286). Among these, N=22 had baseline blood draw for proteomic analysis after the occurrence of a hospitalization record with G12.2. These N=22 comprised the *Clinically Manifest* group in the replication cohort (Table 1). The remaining N=264 ALS cases, whose baseline blood draw occurred before hospitalization record with G12.2, were considered as possible phenoconverters. However, since hospitalization is typically not an early event in the course of ALS, we estimated the approximate phenoconversion time to be two years prior to the G12.2 hospitalization date. After applying this adjustment, N=33 had baseline visits after the approximated time of phenoconversion and were excluded. The remaining N=231 comprised the *Phenoconverter* group in the replication cohort, with a median (range) interval between the baseline visit and the estimated time of phenoconversion being 4.13 (0.05-11.9) years.

Among the N=52,161 participants without a G12.2 record, we further excluded those with any nervous system disease (ICD-10 codes G00–G99; N=15,767), yielding N=36,394 neurologically healthy individuals. Among them, N=35,722 had no pathogenic variants in any ALS genes classified as limited, moderate, strong, or definite by the ClinGen ALS Spectrum Disorders Gene Curation Expert Panel ^42^, or in *HNRNPA1*, *UNC13A*, or *SPTLC1*; they comprised the *Healthy Control* group. The N=38 with either a pathogenic mutation in the SOD1 gene or a *C9orf72* repeat expansion of more than 100 repeats ^43, 44^ comprised the *Pre-symptomatic* group.

For replication analysis, we applied the differential protein expression approach (as described in Step 1 above) to the UKB-PPP cohort, using covariate-matched healthy controls for individuals in the clinically manifest group, with covariates including sex, age, and genotype group. Of the 137 proteins identified in the discovery cohort, 95 were available in UKB due to differences between the Olink Explore 3072 and Olink Explore HT platforms. We plotted the log2fold changes of the overlapping proteins and assessed the correlation between discovery and replication cohorts via Pearson correlation. To analyze the temporal dynamics of proteins in the replication cohort, we applied two-layer generalized additive models (GAMs) instead of two-layer generalized additive mixed models (GAMMs), as the UKB data are cross-sectional rather than longitudinal. Accordingly, individual-level random effects were excluded from the models. In the first-layer GAM, relative protein abundance was modeled as a smooth term of age and covariate sex. The second-layer GAM modeled relative protein abundance against a smooth term of proxy for time-to-phenoconversion, adjusting for sex and genotype group. We then tested our phenoconversion timing estimation models trained on the discovery cohort to baseline protein expression levels in the replication cohort, with covariates including sex, age, and genotype group.

## SUPPLEMENTARY FIGURE LEGENDS

**Supplementary Figure S1. Discovery cohort study schema Supplementary Figure S2. Protein biomarker trajectories (A)** Using data from phenoconverters (pre- and post-conversion) and clinically manifest ALS: Trajectories of 81 protein biomarkers that change pre-symptomatically. For the clinically manifest ALS group, estimated date of symptom onset is used as a proxy for date of phenoconversion. **(B)** Using data from phenoconverters (pre- and post-conversion) only: Trajectories of 74 protein biomarkers that change pre-symptomatically. *(C)* Using only pre-conversion data from phenoconverters: Trajectories of 57 protein biomarkers that change pre-symptomatically. Trajectories are derived from generalized additive mixed model (GAMM) analyses. The Y-axis denotes the log2 fold change in protein relative to age- and sex-matched healthy controls. The dots and connecting lines show the longitudinal data from individual participants. The black curve depicts the estimated mean longitudinal trajectory of protein abundance, adjusting for age and sex; the shaded band denotes the 95% confidence interval around the fitted mean trend. Horizontal red and blue lines indicate the time periods during which protein levels of phenoconverters are, respectively, significantly higher (log2FC>0) or lower (log2FC<0) compared to controls. The numeric labels mark the approximate time at which the significant deviation began to appear.

**Supplementary Figure S3. Protein selection process for discovery cohort** Flowchart explicating stepwise procedures for: identifying proteins that were differentially expressed based on disease state (ALS vs. controls); modeling the longitudinal trajectories of individual protein biomarkers and how they changed before and after ALS phenoconversion; and selecting a core panel of proteins for phenoconversion event prediction, as well as time-to-phenoconversion estimation.

**Supplementary Figure S4. Comparison of phenoconversion event prediction model performance, across different timeframes and machine learning algorithms** Heatmap showing the optimal 5-fold cross-validation AUCs for phenoconversion event prediction, across five timeframes (T = 0.5, 1, 2, 3, and 5 years) and derived from the forward greedy selection method across seven machine learning algorithms. The 137 proteins differentially regulated in ALS vs. controls were included in these analyses. SVM = support vector machine; LASSO = Least Absolute Shrinkage and Selection Operator.

**Supplementary Figure S5. Subject selection process for replication cohort** Flowchart explicating identification of healthy controls, pre-symptomatic carriers, phenoconverters and those with clinically manifest ALS from UK Biobank data. Healthy controls are those with no records of nervous systems diseases and no pathogenic variant in an ALS-associated gene. Pre-symptomatic carriers are those with an *SOD1* mutation or *C9orf72* expansion of >100 repeats, and no hospitalization death record with an ICD-10 code of G12.2. Phenoconverters are those with G12.2 hospitalization date after UK Biobank baseline blood draw; since case identification for the replication cohort relied primarily on hospitalization records, we used 2 years prior to G12.2 hospitalization date as a proxy for phenoconversion. The clinically manifest ALS group includes those with G12.2 hospitalization record prior to baseline blood draw in UK Biobank.

## DATA AVAILABILITY

Requests for access to the discovery cohort Olink proteomic dataset and any associated phenotypic data should be directed to the corresponding author. Provided that the planned research of the requesting institution is fully compliant with the Informed Consent and IRB approvals under which the original samples and phenotypic data have been collected, University of Miami may release said datasets via a Data Transfer Agreement.

## CODE AVAILIABILITY

Code used for analyses in this study is available on https://github.com/ranxm2/ALS_protein_longitudinal.

## ACKNOWLEDGEMENTS

The authors thank the study participants and their families for their altruism, commitment, and contribution to advancing the understanding of pre-symptomatic ALS and efforts toward therapy development and disease prevention; past and current members of the Pre-fALS and CRiALS Biomarker study teams, as well as the CReATe PGB1 study team at the University of Miami, for study coordination and data/biospecimen collection; Dr. Peter M. Andersen at Umea University for genetic testing; and the UK Biobank Pharma Proteomics Project (UKB-PPP) for replication data (accessed under application #847687). The Pre-fALS study has been supported by the Muscular Dystrophy Association (grants 4365 and 172123), ALS Association (grant 2015), NIH (R01 NS105479), ALS Recovery Fund, and Kimmelman Estate. The CReATe PGB1 study was funded through the CReATe Consortium (U54 NS092091), a member of the NIH Rare Diseases Clinical Research Network (RDCRN). The CReATe Biorepository was supported in part by a grant from the ALS Association (grant ID 16-TACL-242).

## AUTHOR CONTRIBUTIONS

- J.W. and M.B. contributed to study conception and design; data acquisition; data analysis and results interpretation; draft of the manuscript; and substantial revision to the manuscript.
- X.R. contributed to data acquisition; data analysis and results interpretation; draft of the manuscript; and substantial revision to the manuscript.
- Z.S.Q. contributed to data analysis and results interpretation; draft of the manuscript; and substantial revision to the manuscript.
- A.M. contributed to study conception and design; data acquisition; and substantial revision to the manuscript.
- V.G., N.C., and P.P. contributed to data acquisition and substantial revision to the manuscript.
- A.L.G., Y.L., E.L., M.F.C., and D.C. contributed to data acquisition.
- J.C.-K. and C.M.L. contributed to substantial revision to the manuscript.
- Collectively, the authors assume responsibility for the completeness and accuracy of the data.

## COMPETING INTERESTS

- V.G. is an employee at Biohaven Pharmaceuticals, and he holds stocks and/or stock options in the company.
- A.M. reports contracts from My Name’5 Doddie Foundation, Target ALS, NIHR UCL Biomedical Research Centre, LifeArc, Medical Research Council, NIH and Motor Neurone Disease Association, Alan Davidson Foundation, Weston Family Foundation, EU H2020 programme; consulting fees from Pfizer, Novartis, LifeArc, Accure, Trace, Neuroscience. Licences to Biogen and ILTOO; and patent numbers, WO2021176044 A1 and WO2024121173A1.
- M.B. reports consulting fees from Alaunos, Alector, Alexion, Amgen, Annexon, Arrowhead, Biogen, Bristol Myers Squibb, Canopy, Cartesian, CorEvitas, Denali, Eli Lilly, Immunovant, Janssen, Merck, Novartis, Prilenia, Roche, Sanofi, Takeda, UCB, uniQure, Voyager, and Woolsey; and is an unpaid member of the Board of Trustees of the ALS Association. The University of Miami has licensed intellectual property to Biogen to support design of the ATLAS study.
- X.R. J.W., Z.S.Q. J.C.-K., A.L.G., Y.L., E.L., M.F.C., D.C., N.C., C.M.L. and P.P. report no competing interests.

## SUPPLEMENTARY TABLES

**Supplementary Table S1.** Count of visits annotated as cases vs. controls for each prediction interval.

| Timeframe | Visit Annotation * | Pre-Symptomatic | Phenoconverters | Total |
| --- | --- | --- | --- | --- |
| T = 0.5 year | Control | 62 | 126 | 188 |
|  | Case | 0 | 37 | 37 |
|  | Total | 62 | 163 | 225 |
| T = 1 year | Control | 62 | 107 | 169 |
|  | Case | 0 | 56 | 56 |
|  | Total | 62 | 163 | 225 |
| T = 2 years | Control | 62 | 86 | 148 |
|  | Case | 0 | 77 | 77 |
|  | Total | 62 | 163 | 225 |
| T = 3 years | Control | 54 | 64 | 118 |
|  | Case | 0 | 99 | 99 |
|  | Total | 54 | 163 | 217 |
| T = 5 years | Control | 36 | 37 | 73 |
|  | Case | 0 | 126 | 126 |
|  | Total | 36 | 163 | 199 |

\* A study visit was annotated as “control” if the participant did not phenoconvert within the specified timeframe, and annotated as “case” if the participant phenoconverted within the specified timeframe.

**Supplementary Table S2.**
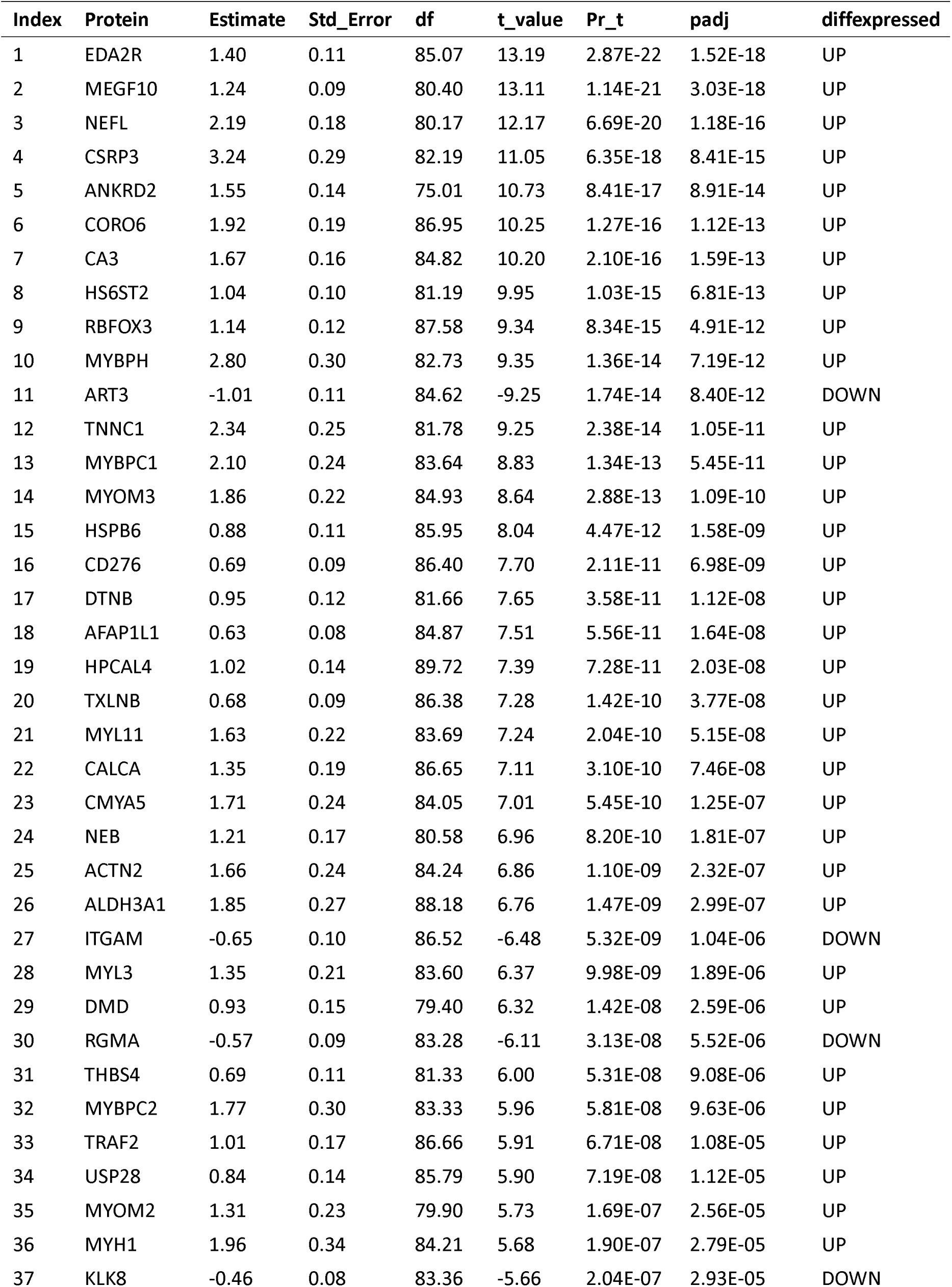
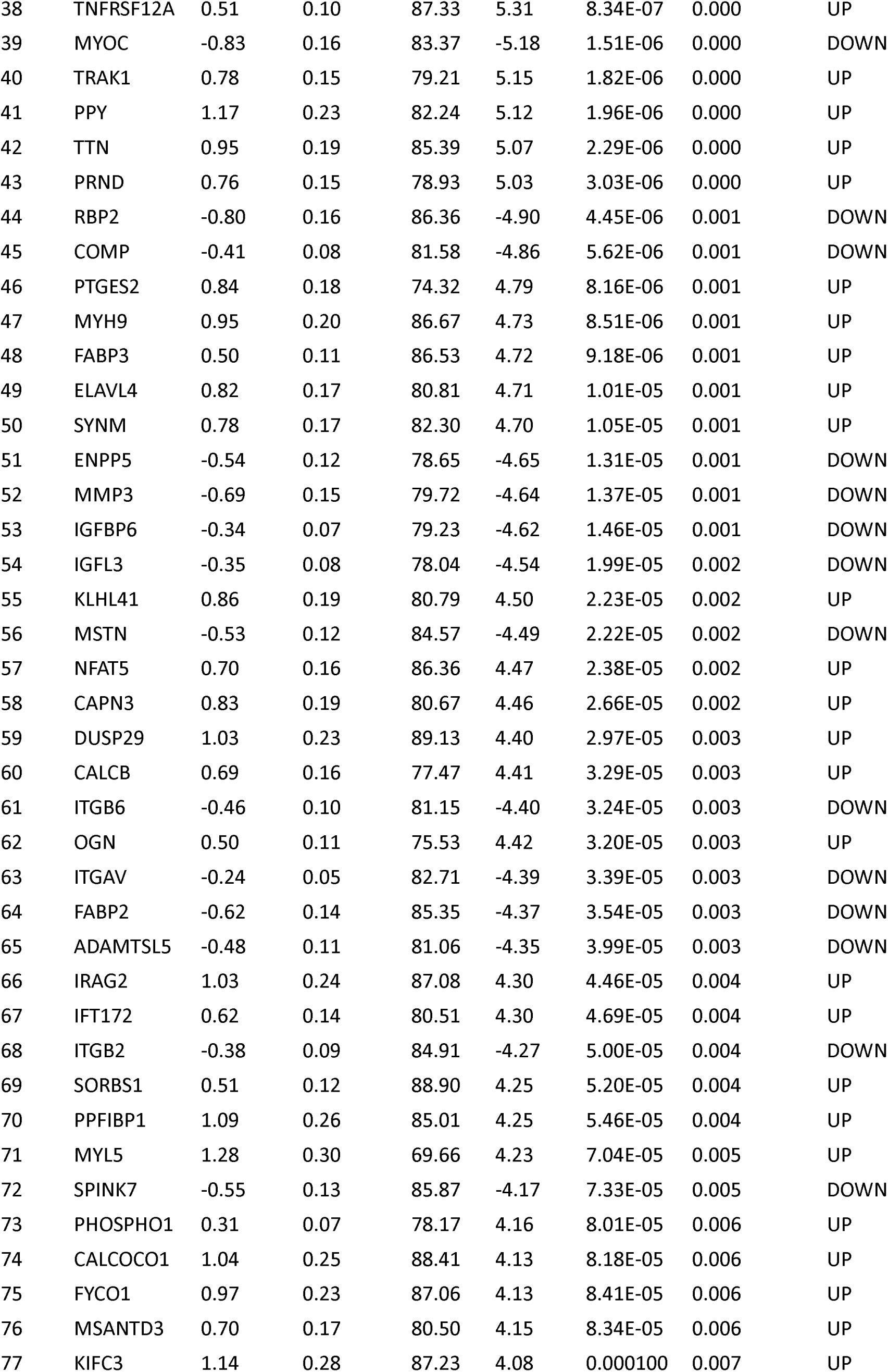
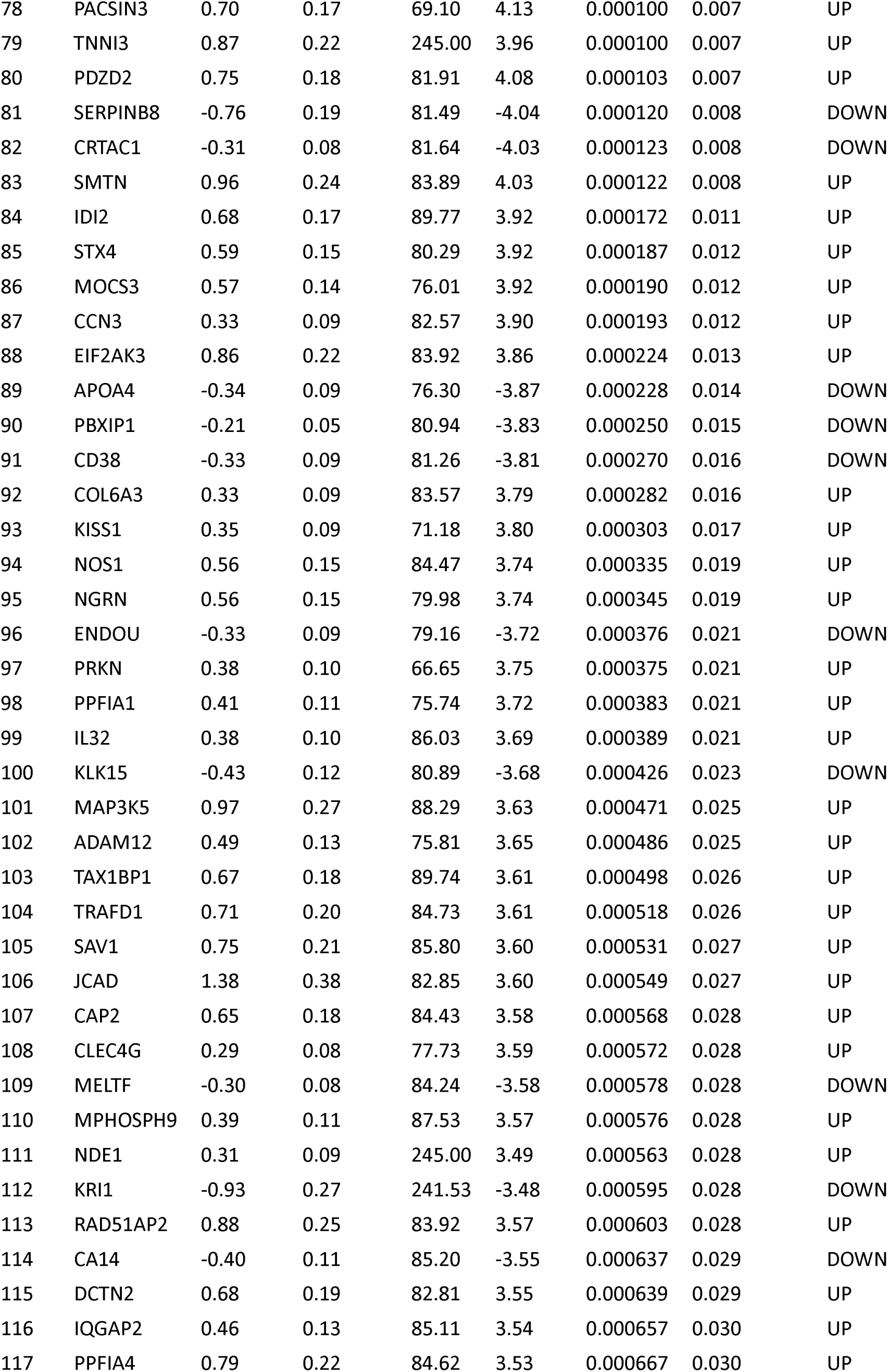
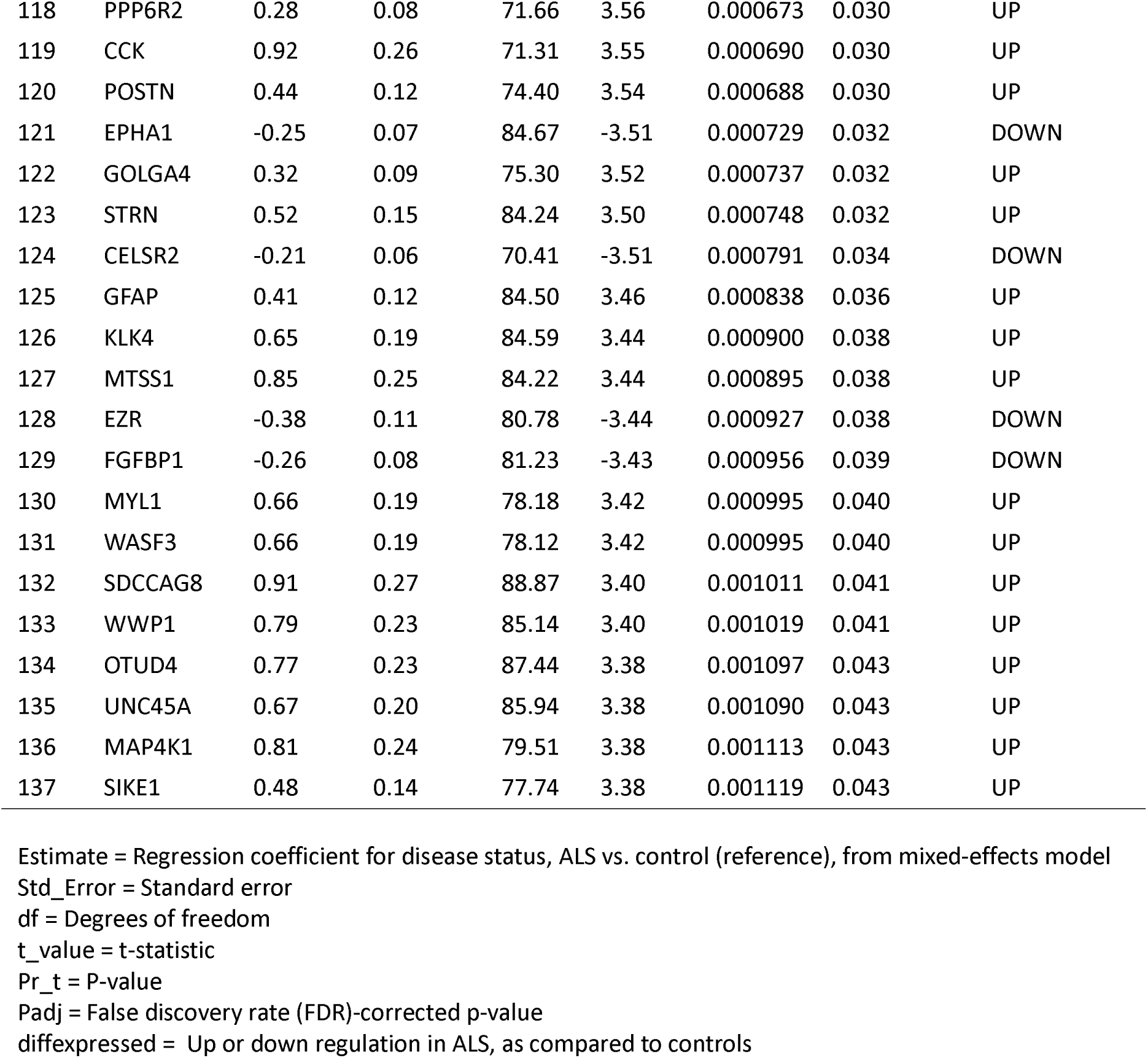
Differential expression proteins in the discovery cohort.

**Supplementary Table S3.**
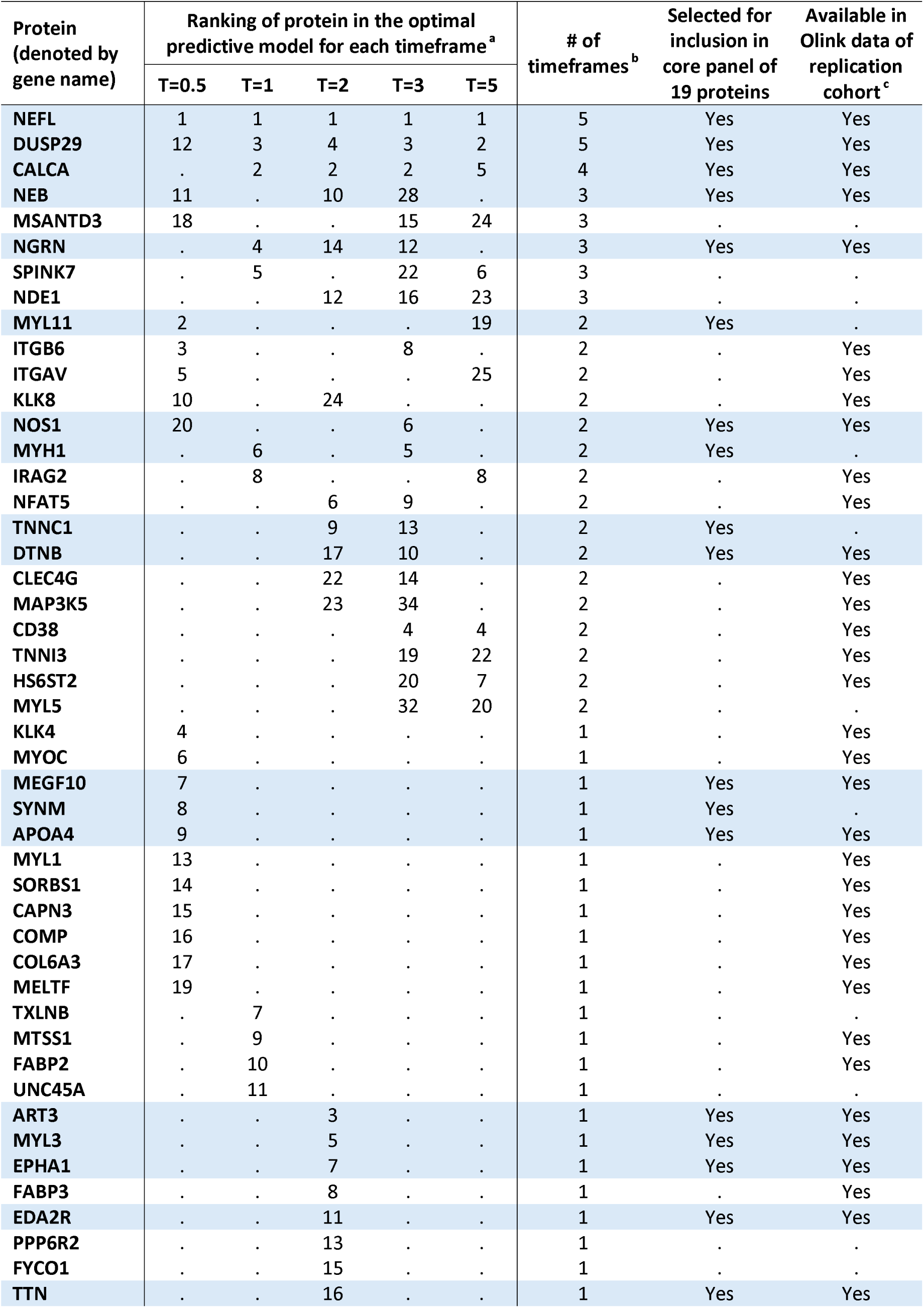
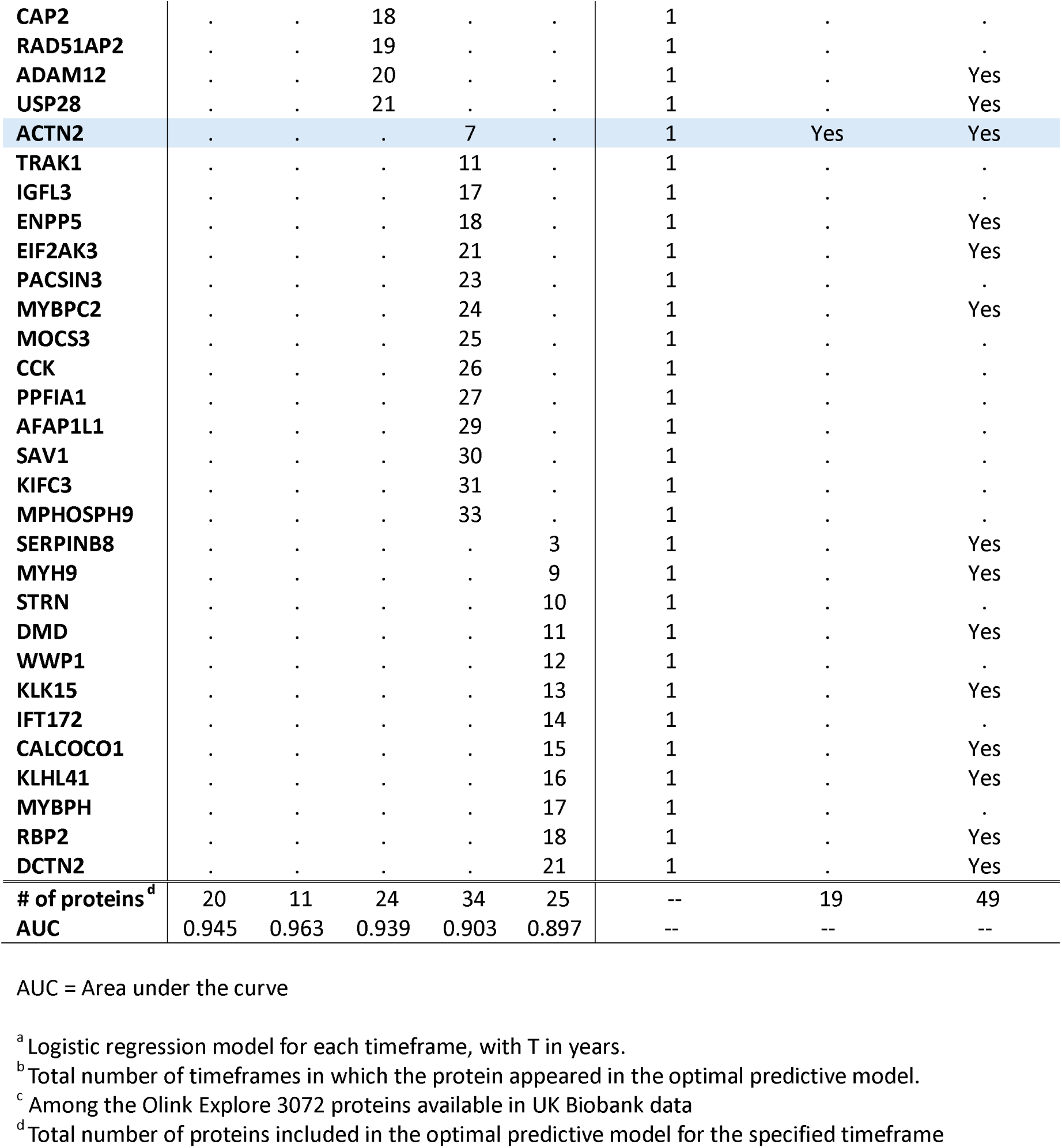
Protein selection for binary phenoconversion prediction across timeframes.

## Notes

### Author Declarations

The University of Miami institutional review board (IRB) gave ethical approval for this work, and all study participants provided written informed consent.

