## Supplementary figures and images for "Predicting Phenoconversion to Clinically Manifest ALS: Results of a Large-Scale Proteomic Study"

### Supplementary Figure S1

#### Figure S1. Discovery Cohort Study Schema

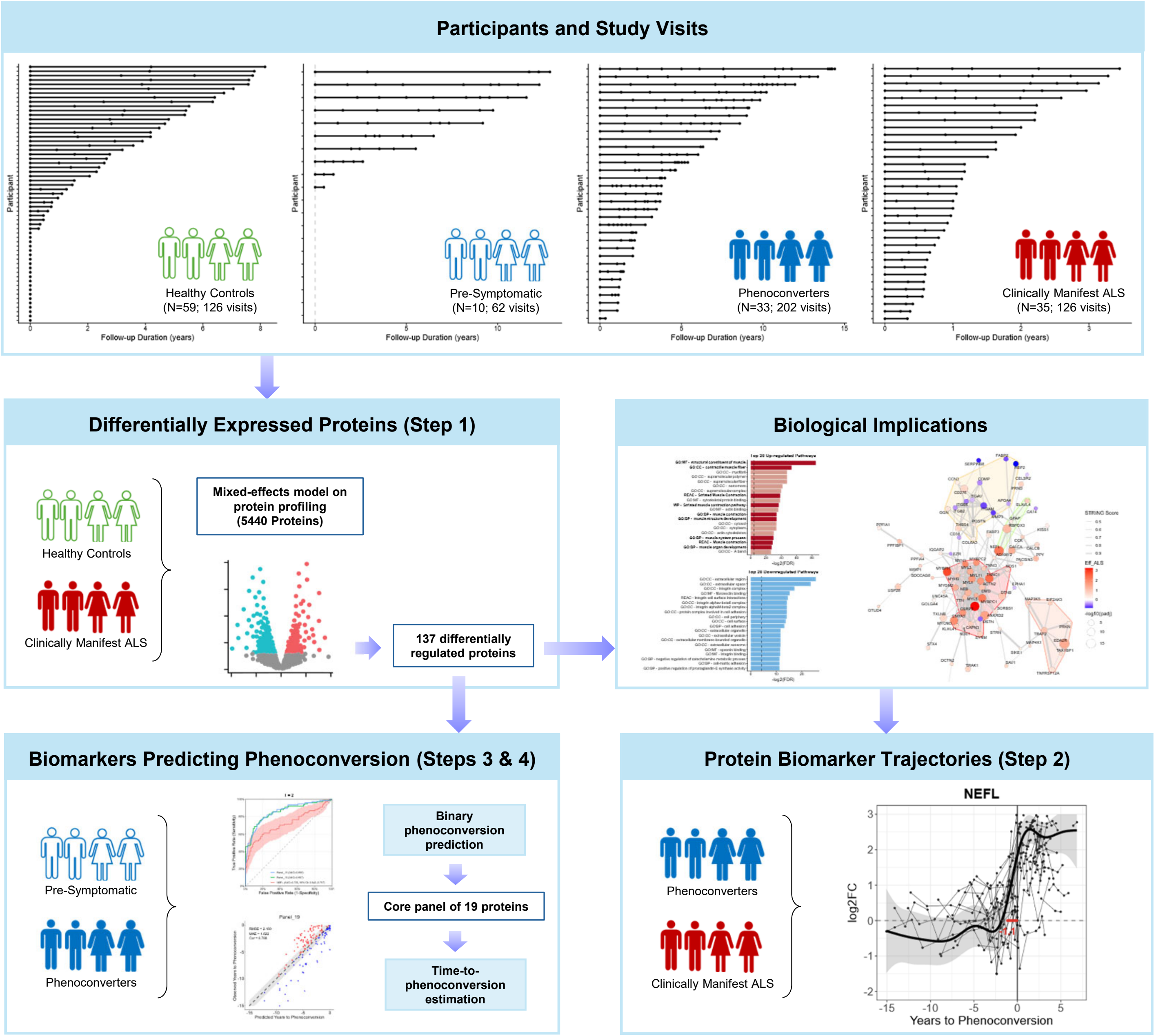

### Supplementary Figure S3

#### Figure S3. Protein Selection Process for Discovery Cohort

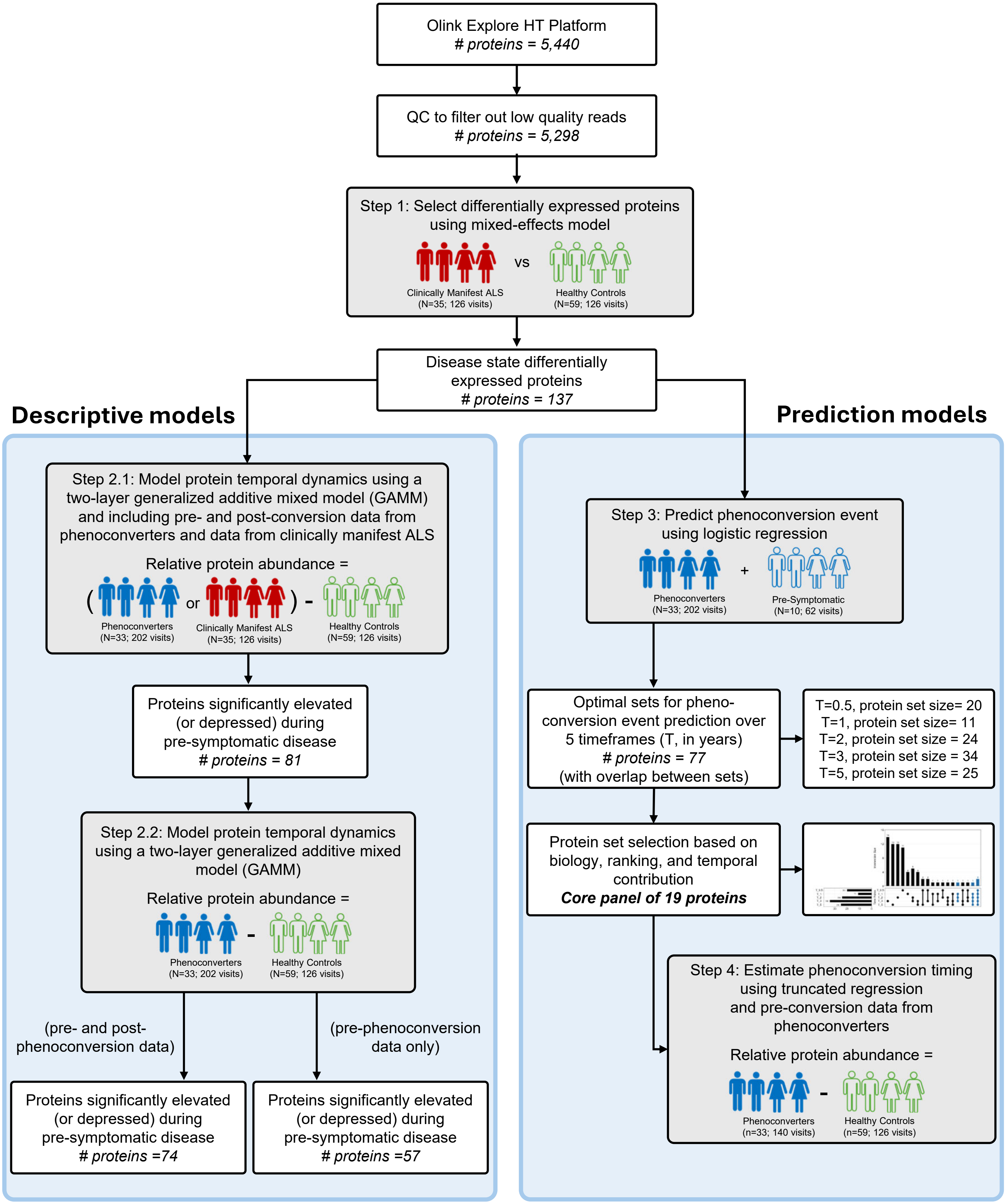

### Supplementary Figure S5

#### Figure S5. Subject Selection Process for Replication Cohort

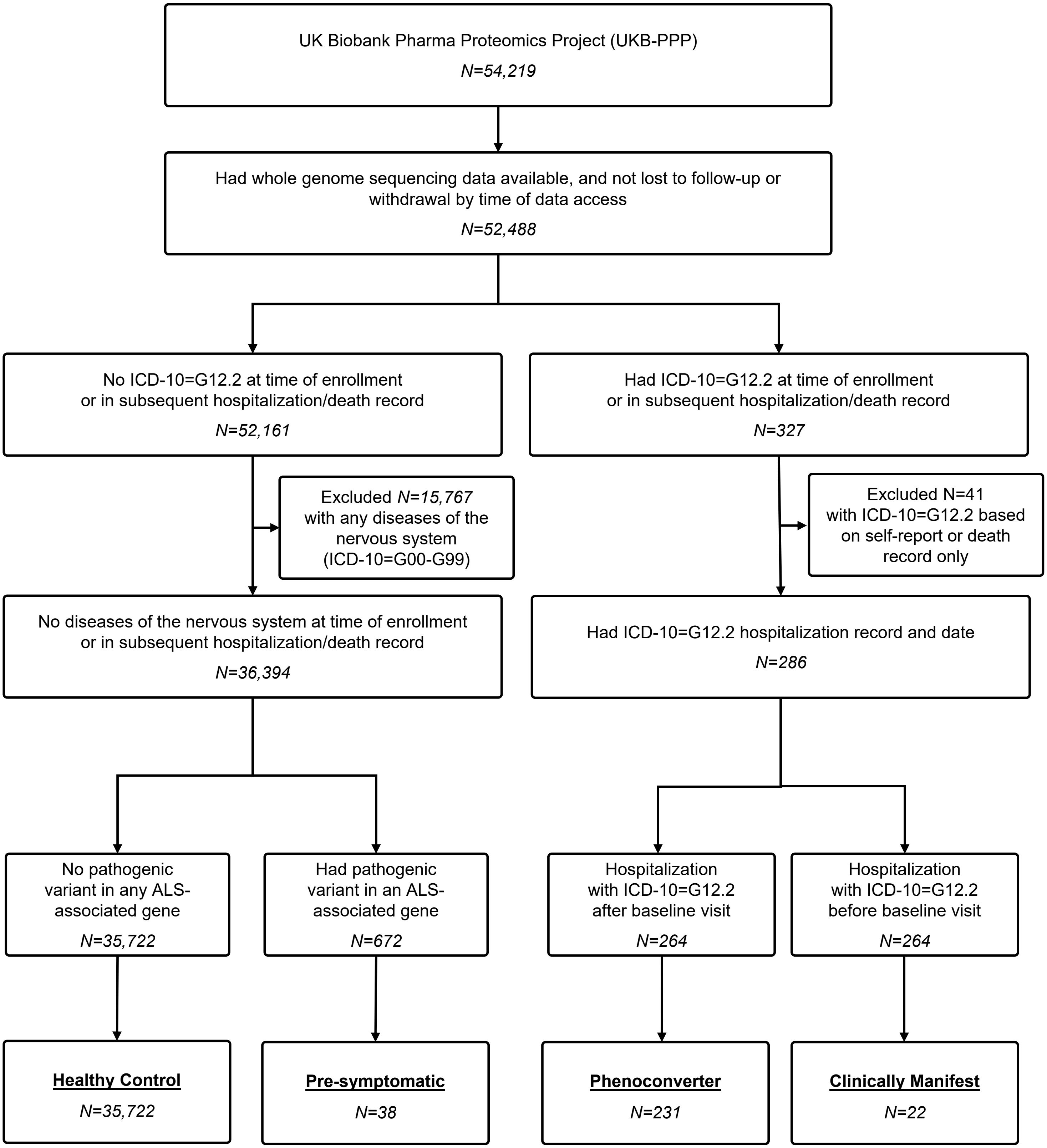
