## Supplementary Figure S2 for "Predicting Phenoconversion to Clinically Manifest ALS: Results of a Large-Scale Proteomic Study"

### Figure S2A. Protein Biomarker Trajectories:
### Using data from phenoconverters (pre- and post-conversion) and clinically manifest ALS

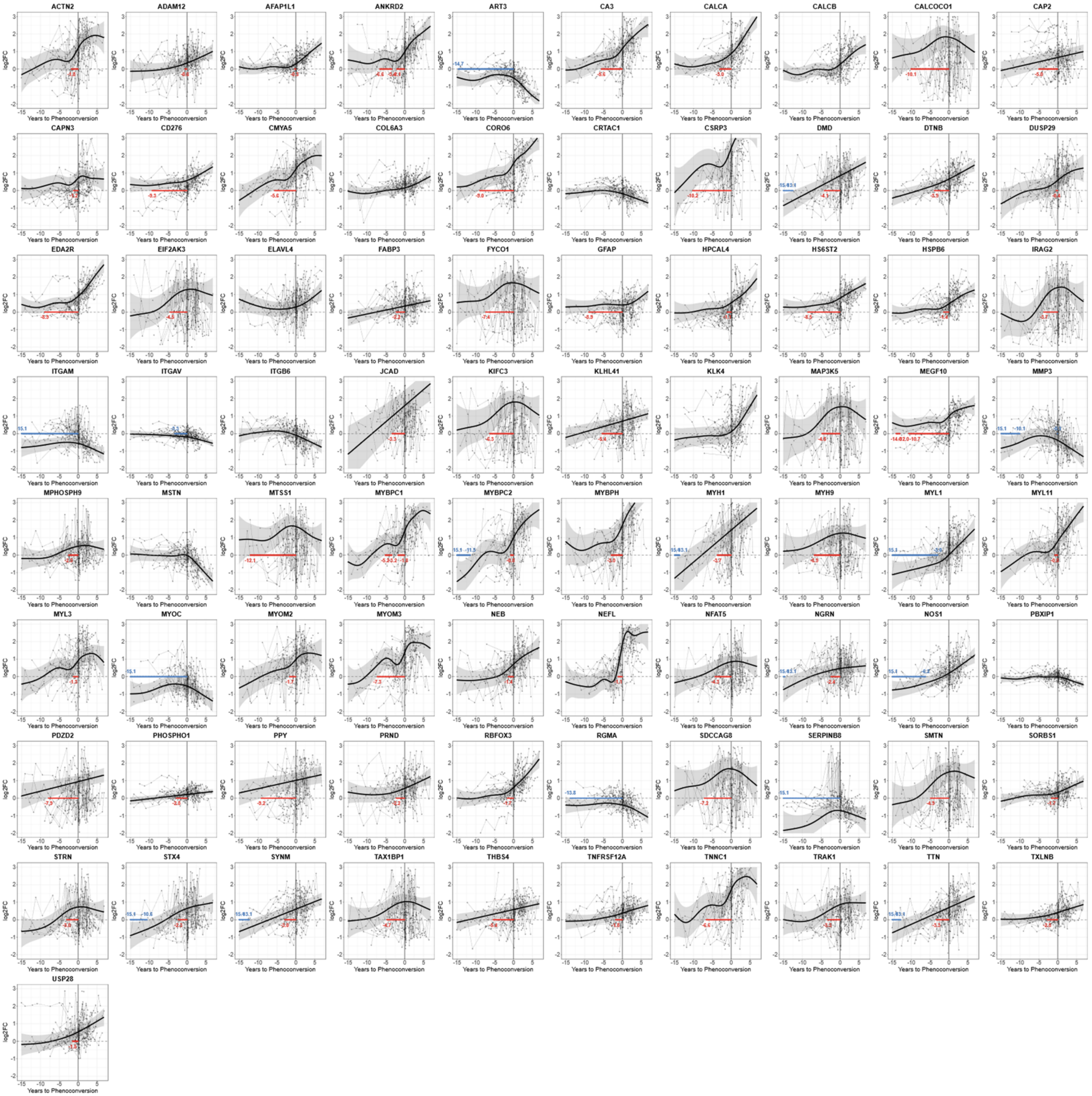

### Figure S2B. Protein Biomarker Trajectories:
### Using data from phenoconverters (pre- and post-conversion) only

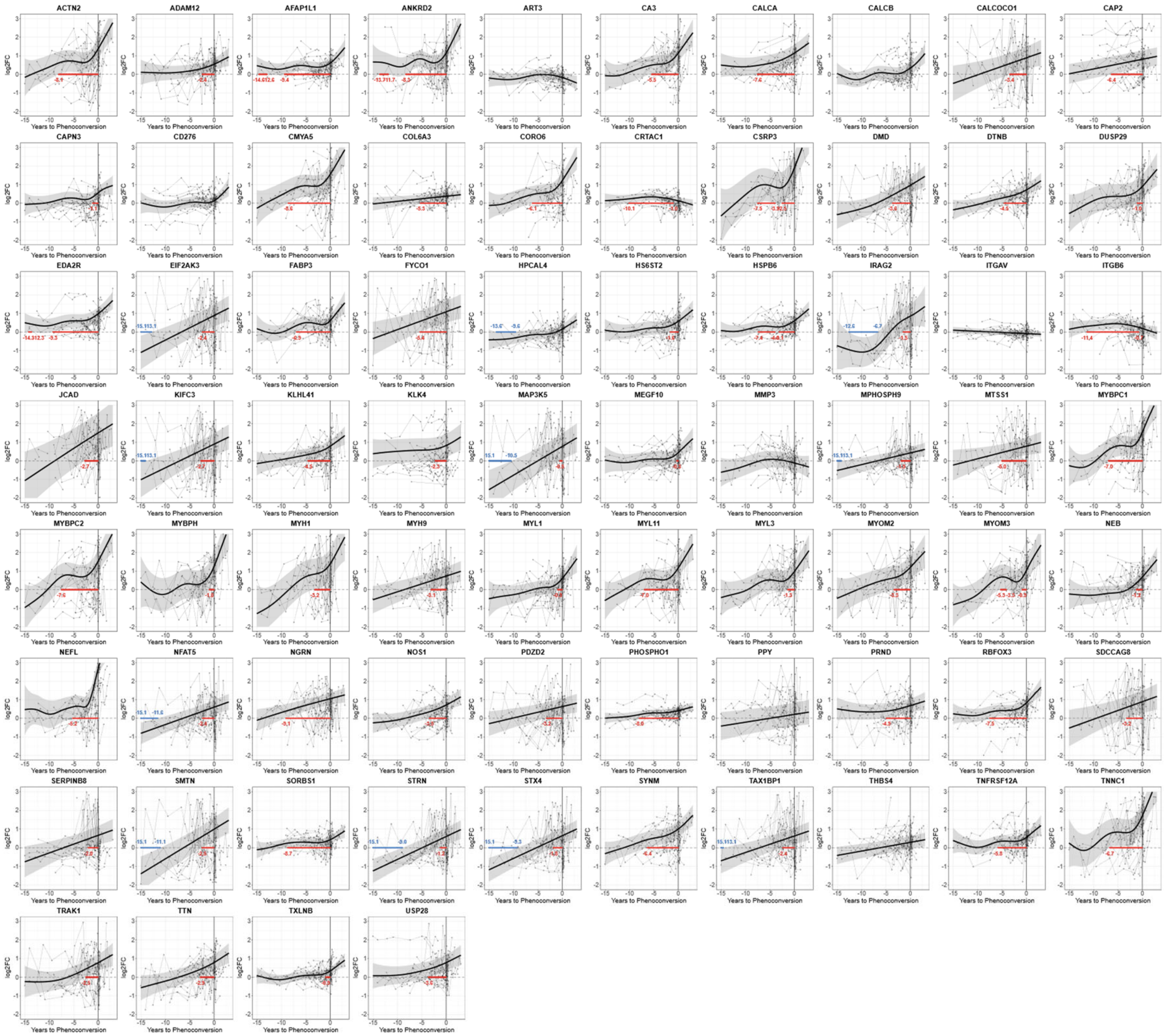

### Figure S2C. Protein Biomarker Trajectories:
### Using only pre-conversion data from phenoconverters

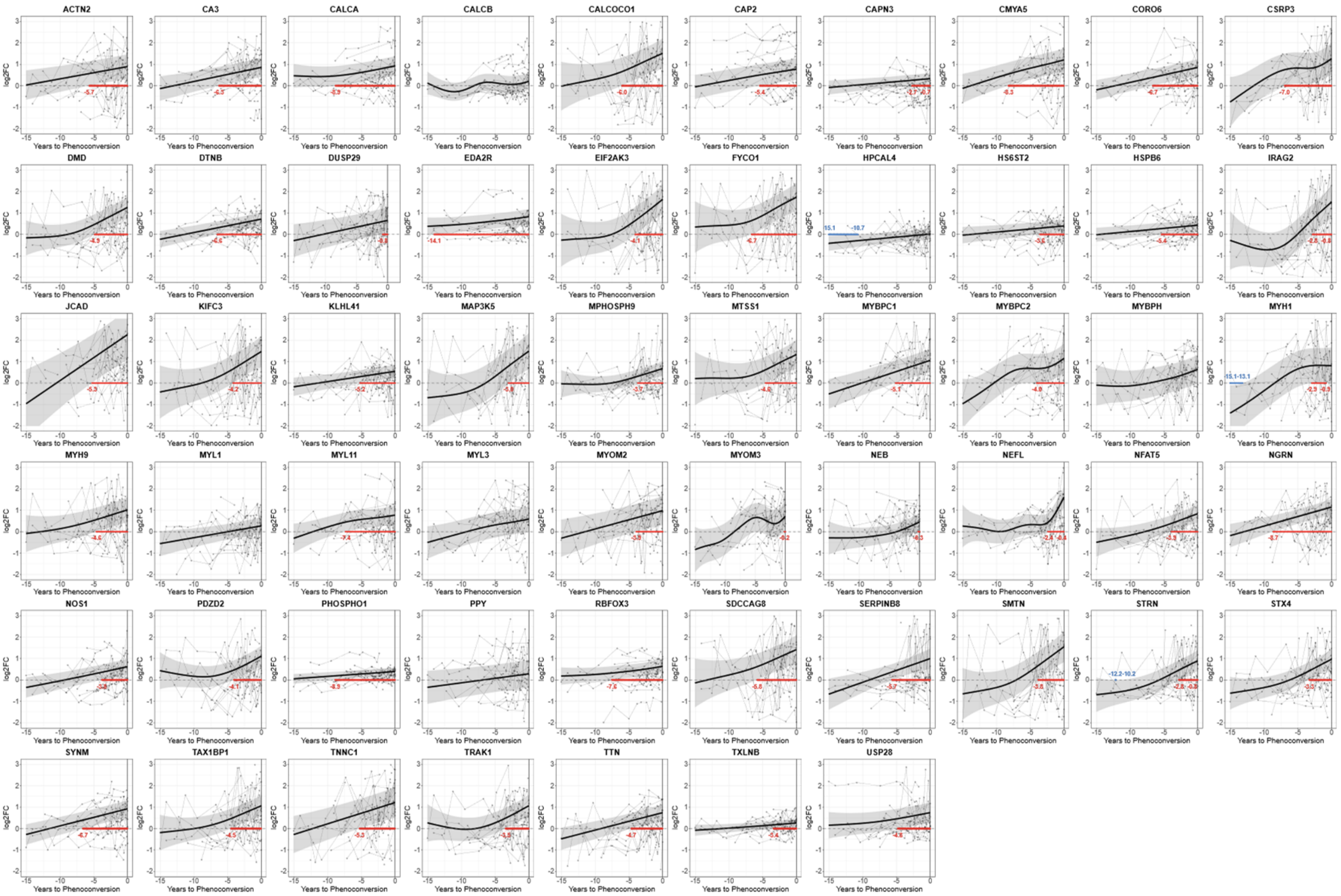
