## Supplementary Figure S4 for "Predicting Phenoconversion to Clinically Manifest ALS: Results of a Large-Scale Proteomic Study"

### Figure S4. Areas Under the Curve for Phenoconversion Event Prediction Using Different Machine Learning Algorithms

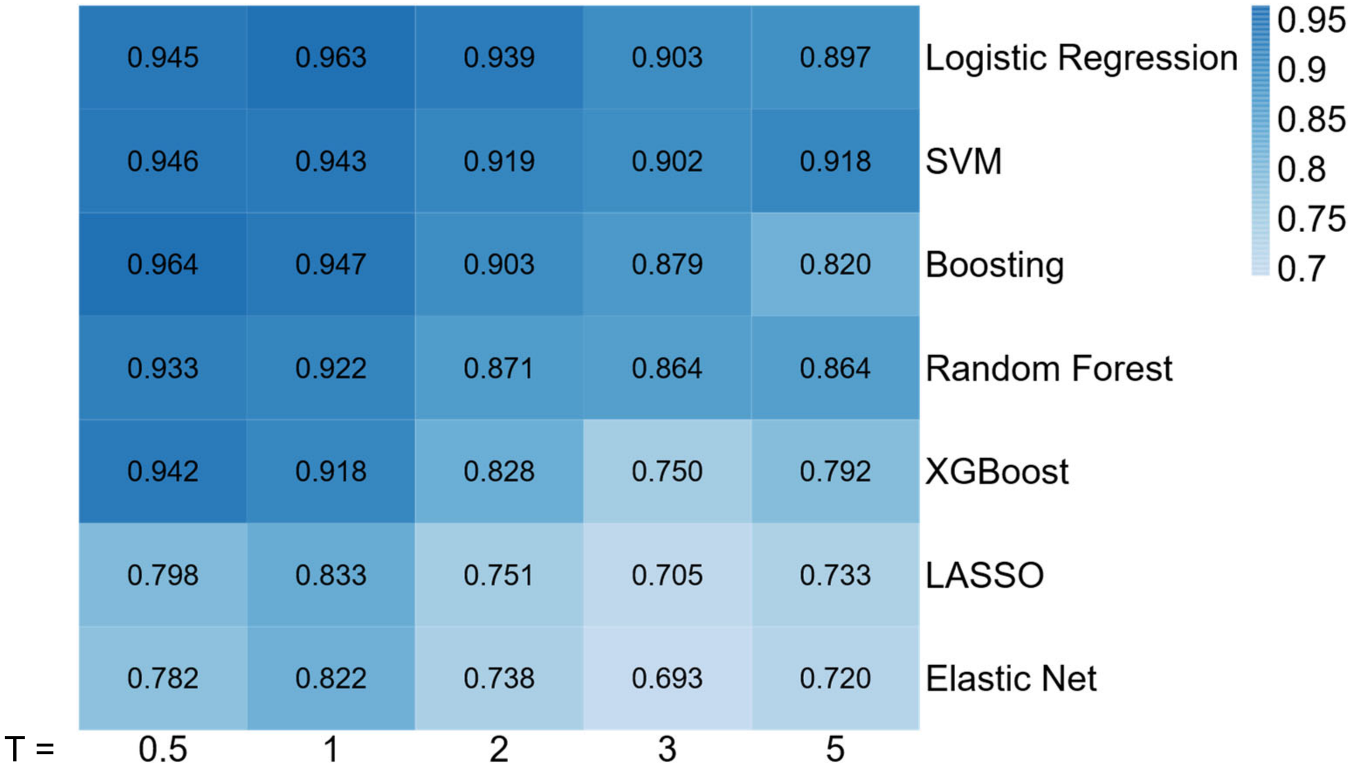
